# Ethnic differences in COVID-19 infection, hospitalisation, and mortality: an OpenSAFELY analysis of 17 million adults in England

**DOI:** 10.1101/2020.09.22.20198754

**Authors:** The OpenSAFELY Collaborative, Rohini Mathur, Christopher T Rentsch, Caroline E Morton, William J Hulme, Anna Schultze, Brian MacKenna, Rosalind Eggo, Krishnan Bhaskaran, Angel YS Wong, Elizabeth J Williamson, Harriet Forbes, Kevin Wing, Helen I McDonald, Chris Bates, Seb Bacon, Alex J Walker, David Evans, Peter Inglesby, Amir Mehrkar, Helen J Curtis, Nicholas J DeVito, Richard Croker, Henry Drysdale, Jonathan Cockburn, John Parry, Frank Hester, Sam Harper, Ian J Douglas, Laurie Tomlinson, Stephen JW Evans, Richard Grieve, David Harrison, Kathy Rowan, Kamlesh Khunti, Nishi Chaturvedi, Liam Smeeth, Ben Goldacre

## Abstract

**Background:** COVID-19 has had a disproportionate impact on ethnic minority populations, both in the UK and internationally. To date, much of the evidence has been derived from studies within single healthcare settings, mainly those hospitalised with COVID-19. Working on behalf of NHS England, the aim of this study was to identify ethnic differences in the risk of COVID-19 infection, hospitalisation and mortality using a large general population cohort in England.

**Methods:** We conducted an observational cohort study using linked primary care records of 17.5 million adults between 1 February 2020 and 3 August 2020. Exposure was self-reported ethnicity collapsed into the 5 and 16 ethnicity categories of the English Census. Multivariable Cox proportional hazards regression was used to identify ethnic differences in the risk of being tested and testing positive for SARS-CoV-2 infection, COVID-19 related intensive care unit (ICU) admission, and COVID-19 mortality, adjusted for socio-demographic factors, clinical co-morbidities, geographic region, care home residency, and household size.

**Results:** A total of 17,510,002 adults were included in the study; 63% white (n=11,030,673), 6% south Asian (n=1,034,337), 2% black (n=344,889), 2% other (n=324,730), 1% mixed (n=172,551), and 26% unknown (n=4,602,822). After adjusting for measured explanatory factors, south Asian, black, and mixed groups were marginally more likely to be tested (south Asian HR 1.08, 95%CI 1.07-1.09; black HR 1.08; 95%CI 1.06-1.09, mixed HR 1.03, 95%CI 1.01-1.05), and substantially more likely to test positive for SARS-CoV-2 compared with white adults (south Asian HR 2.02. 95% CI 1.97-2.07; black HR 1.68, 95%CI 1.61-1.76; mixed HR 1.46, 95%CI 1.36-1.56). The risk of being admitted to ICU for COVID-19 was substantially increased in all ethnic minority groups compared with white adults (south Asian HR 2.22, 95%CI 1.96-2.52; black HR 3.07, 95%CI 2.61-3.61; mixed HR 2.86, 95%CI 2.19-3.75, other HR 2.86, 95%CI 2.31-3.63). Risk of COVID-19 mortality was increased by 25-56% in ethnic minority groups compared with white adults (south Asian HR 1.27, 95%CI 1.17-1.38; black HR 1.55, 95%CI 1.38-1.75; mixed HR 1.40, 95%CI 1.12-1.76; other HR 1.25, 95%CI 1.05-1.49).

We observed heterogeneity of associations after disaggregation into detailed ethnic groupings; Indian and African groups were at higher risk of all outcomes; Pakistani, Bangladeshi and Caribbean groups were less or equally likely to be tested for SARS-CoV-2, but at higher risk of all other outcomes, Chinese groups were less likely to be tested for and test positive for SARS-CoV-2, more likely to be admitted to ICU, and equally likely to die from COVID-19.

**Conclusions:** We found evidence of substantial ethnic inequalities in the risk of testing positive for SARS-CoV-2, ICU admission, and mortality, which persisted after accounting for explanatory factors, including household size. It is likely that some of this excess risk is related to factors not captured in clinical records such as occupation, experiences of structural discrimination, or inequitable access to health and social services. Prioritizing linkage between health, social care, and employment data and engaging with ethnic minority communities to better understand their lived experiences is essential for generating evidence to prevent further widening of inequalities in a timely and actionable manner.

## Background

The risks of COVID-19 infection and outcomes have been reported to be disproportionately increased amongst ethnic minority groups, both in the UK and internationally.^1–7^ It is hypothesized that ethnic differences in COVID-19 infection and outcomes are driven by differences in factors such as living in deprived areas, working in high-exposure or frontline occupations, living in large, multigenerational households, a higher burden of underlying conditions, experiences of discrimination, or access to health and community services.^4,8–12^ As an example, ethnic minority healthcare workers in the UK have experienced higher rates of COVID-19-related death, which has been partly attributed to poorer access to personal protective equipment (PPE and fears around raising concerns about working in unsafe or high-exposure environments.^13,14^

In the UK, the collection of ethnic group data is considered an essential first step towards identifying and actively reducing ethnic inequalities.^15^ Though there is no single universally accepted definition of ethnicity, it serves as an important social construct and surrogate marker for shared exposures or risks for people with similar social, biological, religious, language, and cultural characteristics.^16,17^ The recording of self-reported ethnicity in primary care settings was financially incentivised between 2006-2014, greatly improving the accuracy (completeness, validity and reliability) of these data for clinical care and for research purposes. Recording of ethnic group continues as part of the NHS demographic service, albeit without financial remuneration.^16,18^

To date, studies of COVID-19 have reported findings according to higher-level ethnic groupings, such as, white, south Asian, and black, which may conceal significant heterogeneity.^19,20^ For example, while Bangladeshi and African populations are more likely to live in deprived areas than the general population, Indian and Chinese groups are more likely to live in more affluent areas and experience less material deprivation.^20,21^ Therefore, it is vital to disaggregate broad ethnic groupings to better model the overlapping contributions of health and social factors on COVID-19 infection, severity, and mortality.

Much of the evidence on ethnic differences in COVID-19 has been derived from smaller studies within single healthcare settings, such as those hospitalised with COVID-19.^22–27^ This approach suffers from collider bias, in which factors associated with both COVID-19 infection and hospitalisation, can no longer be explored in an unbiased way, as the study cohort is highly selected and not representative of the general population.^28^ It is also important to look at each stage in the pathway from access to mortality. Furthermore, while previous studies have accounted for health status, social deprivation, or household composition, none have yet explored these factors in conjunction.^29,30^

The aim of this study was to determine ethnic differences across the full pathway for COVID-19, from being tested through to infection, hospitalisation and mortality. Importantly, this study considered the role of socio-demographic factors, clinical co-morbidities, geographic region, care home residency, and household size in both high-level and disaggregated ethnic groups.

## Methods

### Study design and population

We pre-specified and conducted a population-based, observational cohort study using OpenSAFELY, a data analytics platform created on behalf of NHS England to address urgent COVID-19 research questions (https://opensafely.org). OpenSAFELY includes the electronic health record (EHR) data of 24 million people currently registered with primary care practices using TPP SystmOne software, representing approximately 40% of the English population (see supplementary materials for more details).

For this study, primary care data were linked to SARS-CoV-2 antigen testing data from the Second Generation Surveillance System (SGSS), COVID-19 related ICU admissions from the Intensive Care National Audit & Research Centre (ICNARC), and death data from the Office for National Statistics (ONS). The study population comprised all adults, aged 18 years and older, registered with a primary care practice on 1 February 2020. A minimum of twelve months of continuous registration prior to 1 February 2020 was required for inclusion in the study, to ensure that baseline factors were adequately captured. The study period ranged from 1 February 2020 to 3 August 2020.

### Study variables

The primary exposure was self-reported ethnicity as captured on the primary care record. Ethnicity was collapsed into the five and 16 census categories of white (including British, Irish, other white), south Asian (Indian, Pakistani, Bangladeshi, other Asian), black (African, Caribbean, other black), other (Chinese, all other), and mixed (white and Asian, white and African, white and Caribbean, other mixed), and unknown.^31^ Comparisons were reported for the five high-level ethnic groups with the white group as reference, and for the 16 disaggregated ethnic groups, with the white British group as the reference.

Infection-related outcomes included receiving an antigen test for SARS-CoV-2 and testing positive for SARS-CoV-2. COVID-19 disease-related outcomes included being admitted to ICU for COVID-19, and COVID-19-related death (defined as the presence of ICD-10 codes U071 and U072 anywhere in the death certificate).

Demographic characteristics included age, sex, deprivation (defined as quintile of the index of multiple deprivation (IMD)), number of people living in a household (categorised as 1-2 people; 3-5 people; 6-10 people; 11 or more people), care home residency status, number of GP consultations in the 12 months prior to 1 February, and geographic region, defined by the sustainability and transformation partnership (STP) (a National Health Service administrative area).

Clinical covariates included body mass index (BMI), glycated haemoglobin (HbA1c), and blood pressure, defined using the most recent value recorded in the previous ten years. BMI in kg/m^2^ was grouped into six categories using the World Health Organisation classification, which includes adjustments for south Asian ethnicity: underweight (<18 kg/m^2^), normal weight 18.5– 24 (23.5 if south Asian), overweight 25-30 kg/m^2^ (23.6-27.5); obese I 30-34.9 (27.5-32.4); obese II 35-39.9 (32.5-37.4); obese III 40+ (37.5+). Glycated Haemoglobin was grouped into five categories: <6.5%, 6.5-7.4%, 7.5-7.9%, 8-8.9%, >=9%. Blood pressure was grouped into four categories of normal (<120/80), elevated (120-130/80), high stage I (131-140/80-90), and high stage II (>140/90). Smoking status was grouped into current, former and never smokers. Those with missing smoking status were grouped as never smokers.

Clinical comorbidities were considered present at baseline if recorded any time prior to 1 February 2020. These included: hypertension, asthma, chronic respiratory disease, chronic heart disease, type 1 and type 2 diabetes mellitus, cancer, chronic liver disease, stroke, dementia, other chronic neurological diseases, chronic kidney disease (CKD, defined as eGFR<60 ml/min/1.73m^2^), end stage renal failure, common autoimmune diseases (rheumatoid arthritis, systemic lupus erythematosus, or psoriasis), and immunosuppression (HIV, sickle cell disease, organ transplant, asplenia).

Covariates were identified by reviewing literature and through discussions with practising clinicians. Code list creation included clinical and epidemiological review and sign-off by at least two authors. Detailed information on all code lists is openly shared at https://codelists.opensafely.org/ for inspection and reuse.

### Statistical Analysis

Socio-demographic and clinical characteristics at baseline were summarised using descriptive statistics, stratified by ethnic group. Follow-up began on 1 February, 2020 and ended at the earliest of experiencing an outcome of interest, or death, de-registration from a TPP primary care practice, or the censoring date for the dataset for capturing the outcome of interest (between 30 July and 3 August, 2020).

Multivariable Cox proportional hazards regression was used to estimate ethnic differences in the risk of each outcome. All analyses were adjusted for the pre-specified covariates listed above. These included age (using restricted cubic splines), sex, deprivation quintile, all pre-specified clinical co-morbidities, BMI, HbA1c, and blood pressure, number of primary care consultations in the prior twelve months, household size, care home residency, and stratification by STP region.

### Sensitivity Analysis

Two sensitivity analyses were conducted. Firstly, in addition to estimating ethnic differences in the risk of testing positive for SARS-CoV-2 in the general population, ethnic differences in the odds of testing positive amongst those ever tested were estimated using multivariable logistic regression. Secondly, we estimated ethnic differences in non-COVID-related death for comparison with COVID-19-related death.

### Software and Reproducibility

Data management was performed using Python 3.8 and SQL, and analysis was carried out using Stata 16. The pre-specified protocol and code for data management and analysis are archived online at https://github.com/opensafely/ethnicity-covid-research.

## Results

From a total of 23,600,617 people actively contributing to the OpenSAFELY platform on 1 February, 2020, 17,510,002 adults, aged 18 or over, with at least twelve months of prior registration were included in the study (Figure 1).

**Figure 1.**
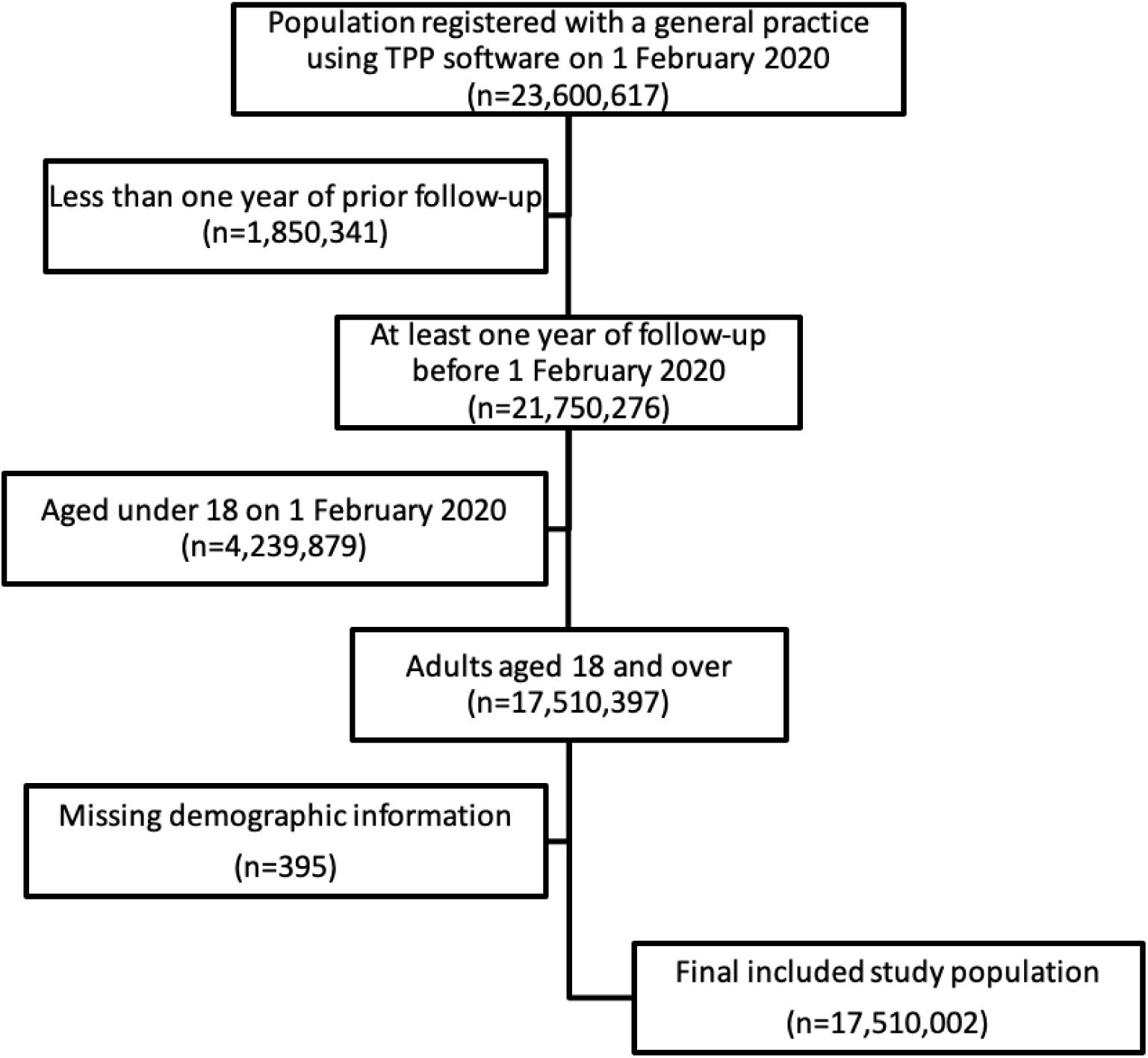
Population inclusion Flowchart.

The ethnic breakdown of the cohort was 63% white (n=11,030,673), 6% south Asian (n=1,034,337), 2% black (n=344,889), 2% other (n=324,730), 1% mixed (n=172,551), and 26% unknown (n=4,602,822) (Table 1). The 16-category breakdown of ethnicity was 54.8% White British, 0.5% Irish, 7.7% other white, 0.5% Indian, 1.1% Pakistani, 0.4% Bangladeshi, 0.6% other Asian, 1.3% African, 1.0% Caribbean, 0.4% other Black, and 0.6% Chinese (Table S1).

**Table 1.**
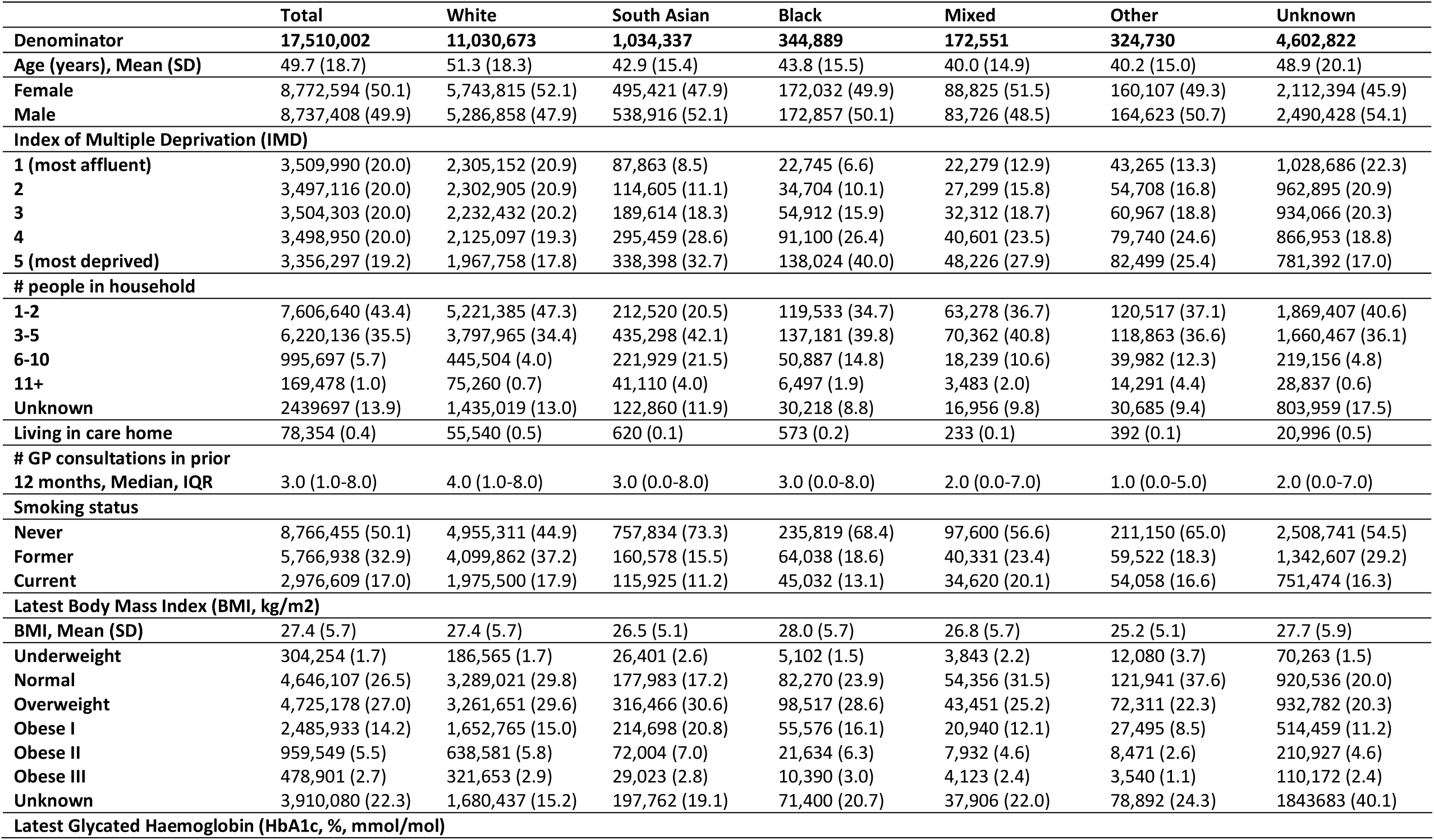
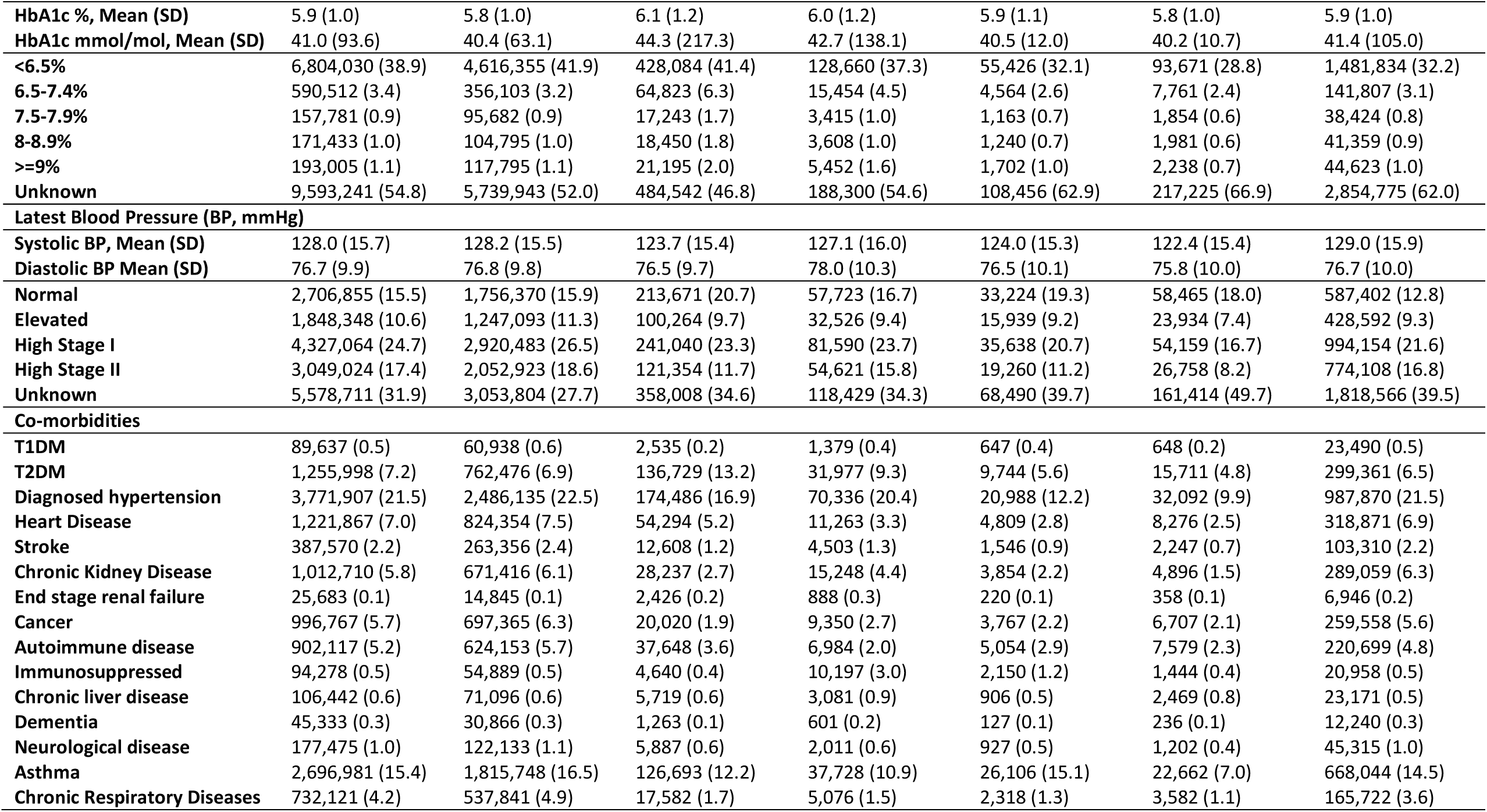
Baseline Characteristics by ethnic group (5 categories)

Compared with the white population, ethnic minority groups were, on average, ten years younger, over-represented in deprived neighbourhoods and large households, and under-represented in care home populations. The prevalence of type 2 diabetes was higher in south Asian groups compared with white groups (13.2% vs. 6.9%; Table 1). When examined in greater details, Bangladeshi and Chinese groups were youngest and Pakistani groups had the highest proportion of individuals in residing in deprived neighbourhoods and households of six or more people. Bangladeshi and Caribbean groups had the highest prevalence of type 2 diabetes (Table S1).

Those with unknown ethnicity were more similar to the White group with respect to age, deprivation, care home residency, household size, BMI, and prevalence of co-morbidities. Median number of consultations in the prior 12 months was comparable to the mixed ethnic group (Tables 1, S1).

### SARS-CoV-2 testing and positive rates in the general population

Between 1 February and 3 August 2020, 8.9% of the study population received an antigen test for active SARS-CoV-2 infection (n=1,552,521), and 0.5% tested positive (n=82,473) (Table S2).

After accounting for all measured explanatory factors, south Asian, black, and mixed groups were more likely to be tested for SARS-CoV-2 (south Asian HR 1.08, 95% CI 1.07-1.09; black HR 1.08 95% CI 1.06-1.09; mixed HR 1.03, 95% CI 1.01-1.05), and more likely to have a positive test result recorded (south Asian HR 2.02 95% CI 1.97-2.07; black HR 1.68, 95% CI 1.61-1.76; mixed HR 1.46, 95% CI 1.36-1.56; Figure 2)

**Figure 2.**
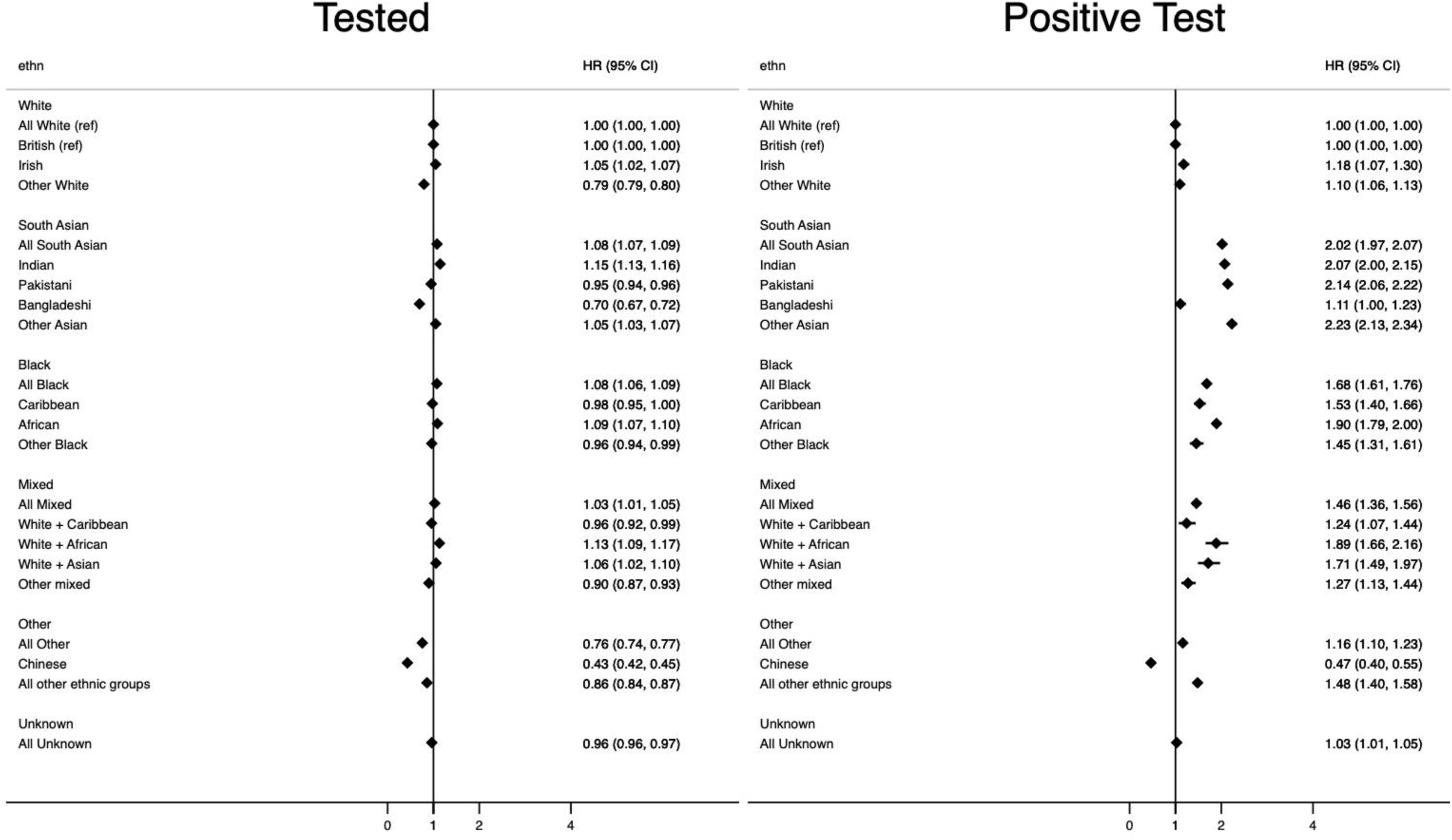
Ethnic differences in being tested for and testing positive for SARS-CoV-2 infection. ∗All White is the reference category for comparison of ethnicity in 5 categories. British is the reference category for comparison of ethnicity in 16 categories. ∗models adjust for age, sex, deprivation quintile, all pre-specified clinical co-morbidities, categories of BMI, HbA1c, and systolic and diastolic blood pressure, number of primary care consultations in the 12 months prior, household size, care home residency, and stratification by STP region.

When broken down into the 16 categories of ethnicity, the likelihood of being tested was higher for Irish (HR 1.05, 95% CI 1.02-1.07), Indian (HR 1.15, 95% CI 1.13-1.16) and African groups (HR 1.09, 95% CI 1.07-1.10), and lower for Other white (HR 0.79, 95%CI 079-0.80), Pakistani (HR 0.95, 95% CI 0.94-0.96), Bangladeshi (HR 0.70, 95% CI 0.67-0.72), and Chinese groups (HR 0.43, 95% CI 0.42-0.45) compared with the white British group. The likelihood of having a positive test result recorded was higher in all ethnic minority groups compared with the white British group, except for the Bangladeshi group for whom risks were similar (HR 1.11, 95%CI 1.00-1.23), and for the Chinese group, for whom risks were lower (HR 0.47, 95% CI 0.40-0.55; Figure 2).

There was evidence of a small difference between those of unknown and white group with respect to being tested for SARS-CoV-2 (HR 0.96, 95%CI 0.96-0.97), and testing positive (HR 1.03, 95%CI 1.01-1.05; Figure 2).

### Severe COVID-19 outcomes

Of the total study population, <0.1% were admitted to ICU for COVID-19 (n=3,118), and 0.1% had a COVID-19-related death (n=15,627) (Table S2).

After accounting for all measured explanatory factors, the risk of being admitted to ICU for COVID-19 was higher in the south Asian (HR 2.22, 95% CI 1.96-2.52) Black (HR 3.07, 95% CI 2.61-3.61) mixed (HR 3.2.86, 95% CI 2.19-3.75), and other (HR 2.86, 95% CI 2.31-3.53) ethnic groups compared with the white group. This pattern remained consistent for all ethnic subgroups, including Chinese (HR 2.44, 95% CI 1.53-3.89); Figure 3.

**Figure 3.**
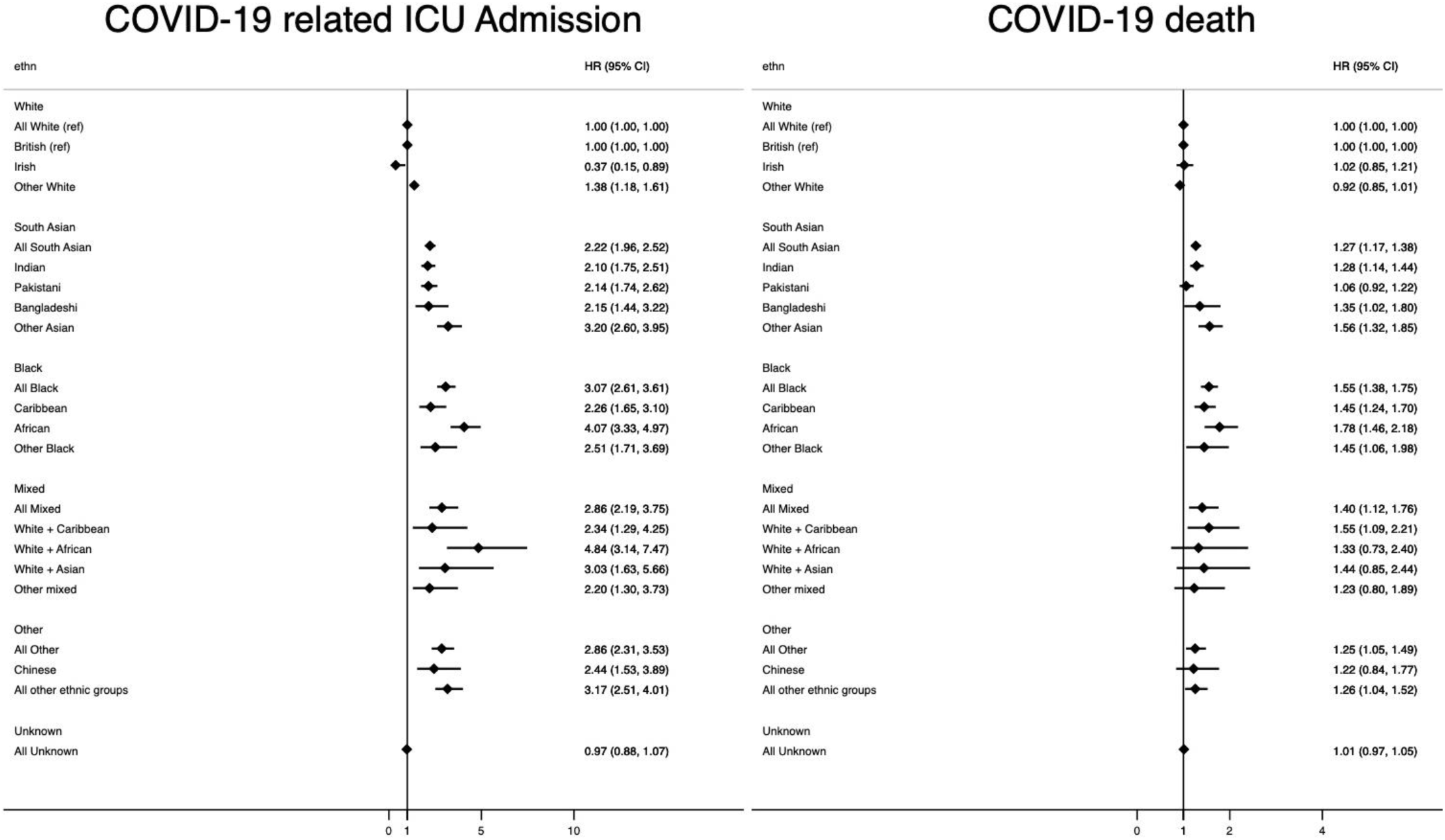
Ethnic differences in risk of COVID-19 related ICU admission and mortality. ∗All White is the reference category for comparison of ethnicity in 5 categories. British is the reference category for comparison of ethnicity in 16 categories. ∗models adjust for age, sex, deprivation quintile, all pre-specified clinical co-morbidities, categories of BMI, HbA1c, and systolic and diastolic blood pressure, number of primary care consultations in the 12 months prior, household size, care home residency, and stratification by STP region.

The risk of COVID-19 death was increased by 27% in the south Asian group (HR 1.27, 95% CI 1.17-1.38), 56% in the black group (HR 1.56, 95% CI 1.38-1.75), 40% in the mixed group (HR 1.40, 95% CI 1.12-1.76), and 25% in the other ethnic group (HR 1.25, 95% CI 1.05-1.49). When broken down further, the relative risk of COVID-19 death was highest in the African (HR 1.78, 95%CI 1.46-2.18), mixed white + Caribbean (HR 1.55, 95%CI 1.09-2.21), and other Asian groups (HR 1.56, 95% CI 1.32-1.85), and equivalent between white and Chinese groups (HR 1.22, 95%CI 0,80-1.89; Figure 3).

No differences in the risk of COVID-19 related ICU admission or death were apparent for those in the unknown ethnic group compared with the white group (ICU HR 0.97, 95%CI 0.88-1.07, COVID-19 death HR 1.01, 95%CI 0.97-1.05; Figure 3).

### Sensitivity Analyses

Ethnic group differences in the odds of testing positive remained unchanged within the subset of individuals ever tested for SARS-CoV-2 for most groups. However, while the risk of testing positive was lower among the Chinese group in the general population, no differences in the odds of having a positive test recorded were observed after restricting to the population tested (OR 1.09, 95% CI 0.92-1.29, Figure S1). A total of 84,872 non-COVID related deaths occurred over the study period. The risk of non-COVID-related death was reduced by 13-28% in all non-white ethnic groups compared with the white group; south Asian HR 0.82, (95%CI 0.78-0.87), black HR 0.87 (95%CI 0.80-0.94), mixed HR 0.79 (95%CI 0.64-0.80), other HR 0.72 (95%CI 0.64-0.80; Table S4, S5).

## Discussion

### Summary

We found clinically important ethnic differences for testing positive for SARS-CoV-2 infection, for COVID-19 ICU admission and for COVID-19 related mortality in the largest European study to date, drawing on the full population-based clinical records for 17 million adults. We observed that ethnic differences persisted even after accounting for key explanatory factors such as socio-demographic factors, clinical co-morbidities, geographic region, care home residency, and household size.

Compared with the white British ethnic group, Indian, and African groups were at higher risk of all outcomes studied; namely, being tested for and testing positive for SARS-CoV-2, being admitted to ICU for COVID-19, and having a COVID-19-related death. Pakistani and Bangladeshi groups were less likely to be tested for SARS-CoV-2, but more likely to test positive, be admitted to ICU and die from COVID-19. Caribbean groups were equally likely to receive a test for SARS-CoV-2 but had higher risk of all other outcomes. Chinese groups were less likely to be tested for and test positive for SARS-CoV-2, more likely to be admitted to ICU, and equally likely to die from COVID-19. Irish and other White groups were more likely to test positive for SARS-CoV-2, but equally likely to die from COVID-19 compared with the British group.

## Strengths and limitations

The greatest strength of this study the sample size and the use of raw event-level data that was not aggregated before analysis. This large, population-based cohort captured high quality clinical data, across a range of healthcare settings, and linked COVID-19 data sources which allowed us to provide insight into disparities between ethnic groups at different stages of the COVID-19 pathway prior to mortality. We were able to report findings according to self-reported ethnicity in 16 groups whereas many other UK based studies have aggregated self-reported ethnicity into higher-level groups due to small numbers. Finally, we reported differences in outcomes using a population-based sample, which allowed us to overcome issues faced by studies limited to individuals with evidence of SARS-CoV-19 infection, or hospitalized with COVID-19, where populations under study do not represent the true general population at risk.^28^

We also recognise some limitations. Ethnicity data were missing for 26% of the study population. Previous work in OpenSAFELY, however, has reported no differences in associations between ethnic group and COVID-19 death after accounting for missing data using multiple imputation.^2,32^ Here, we found that clinical and demographic characteristics of the unknown ethnic group and risk of outcomes most closely mirrored the white ethnic group.

Our inability to capture all explanatory factors is likely to have impacted our observed findings. For example, factors associated with ethnicity (such as ancestry, occupation, experiences of racism or discrimination, and behaviour) were unmeasured, while other factors, such as severity and progression of comorbid conditions were not measured in detail. We were unable to estimate household size for 13% of our population, as these households had invalid address information. We may have underestimated household size for homes including people registered at non-TPP primary care practices, and we may have over-estimated household size for individuals living in large apartment blocks, or for people who have not updated their address after moving, making them appear to reside in their old address alongside the current occupants. In recognition of these limitations, we grouped household size into four levels rather than considering it as a continuous measure.

We also note a general limitation with studies using SARS-CoV-2 test data as an outcome in English primary care. Due to the selection of people who have the opportunity to be tested, it is currently impossible to disentangle whether observed differences in outcomes are due to ethnicity or due to having received a test (collider bias).^28^ Two possible selection mechanisms are firstly, that people who have symptoms are more likely to be tested and test positive, and secondly, people residing in high-transmission geographic areas or working in high-exposure professions may have more opportunity to be tested and test positive, even if asymptomatic. In the absence of population-representative testing, we did not attempt to examine ethnic group differences in the risk of COVID-19 outcomes in individuals with evidence of infection or disease as we do not view test results as a reliable way of identifying a representative infected population. Over time, as population-based testing becomes more widespread, understanding ethnic differences in prognosis for people infected with SARS-CoV-2 will be important to determine.

## Findings in Context

Our results mirror those of a recent USA report of 55 million patients in the EPIC database, which found that Hispanic, black, and Asian groups had equivalent rates of testing, but higher rates of infection, hospitalization and death compared to the white population after accounting for socio-demographic characteristics and underlying health conditions.^33^ Though yet to be peer-reviewed, these parallel findings suggest that potential mechanisms underpinning ethnic differences in COVID-19 outcomes in the UK, such as structural discrimination and occupational risk, may be common in other settings.

Data from the 2011 Census shows that south Asian and black groups are more likely to live in large multi-generational households than white groups, and in over-crowded spaces.

These factors may both influence exposure to infection and ability to isolate if in a high-risk or vulnerable population group.^34,35^ By accounting for household size and care home residency, we have shown that these factors contribute to, but do not fully explain differences by ethnic group.

Exploring differences between the detailed ethnic groups revealed important heterogeneity. For example, south Asian groups, as a whole, were more likely to be tested for COVID-19, but this was true only for the Indian group, with Pakistani and Bangladeshi groups less likely to receive a test. This difference may be related to the fact that Indian groups make up 14% of all doctors (compared with 3% of the general population), and opportunities for testing were much higher for healthcare workers than the general population, particularly in the early phases of the UK epidemic.^36^ Lower rates of testing in Bangladeshi and Pakistani populations may also be related to poorer health literacy in these groups, lack of tailored or accessible health communications, or lack of access to testing facilities.^37^ Our finding that Chinese groups were less likely to be tested for SARS-CoV-2, but equally likely to test positive amongst those ever receiving a test, may relate to differences in health seeking behaviour, or our inability to capture test results for those who were tested outside the UK (for example, if individuals left the country when travel restrictions were imposed).^38^

The ONS has reported “…Of the 17 specific occupations among men in England and Wales found to have higher rates of death involving COVID-19, data from the Annual Population Survey (APS) show that 11 of these have statistically significantly higher proportions of workers from Black and Asian ethnic backgrounds…”. Their analysis included deaths registered between 9 March and 25 May 2020 in adults aged 20 to 64 years in England and Wales and they “adjusted for age, but not for other factors such as ethnic group and place of residence”. Our analyses did not have occupation but did include age, area of residence, medical history, and household size. Our study suggests that findings of increased risks among ethnic minority groups are likely to have, at least, a partial explanation associated with occupation.^39^ Prioritizing linkage between health, social and employment data will be essential in building a complete picture of ethnic group differences in COVID-19 risk and outcomes.

The COVID-19 pandemic has highlighted the urgent need for data disaggregated by ethnic group. The heterogeneity of associations in this study suggest that analyses using detailed ethnic groupings should be the standard where possible. Our study has shown that there are major differences in COVID-19 outcome by ethnicity, not all of which can be explained by the health or demographic factors that we can measure. There are unexplained differences which must be tackled to reduce the health inequality that COVID-19 has highlighted in the UK. Awareness of these inequalities is a necessary first step to policy reform around improving access to testing and healthcare, including admission to ICU, particularly for black and south Asian groups who we found to be at highest risk for COVID-related death.

National data from England and Scotland have shown that most ethnic minority groups have both better overall health and lower rates of all-cause mortality than white groups.^40,41^ We were able to confirm this pattern in our sensitivity analyses. Thus our findings of disparities in COVID-19 related infection and outcomes are particularly concerning.^40,41^

Improving the quality and completeness of ethnicity data across health and administrative datasets is essential for building a complete picture of ethnic group disparities. Despite historic financial investment, the recording of ethnicity in GP records remains incomplete. Furthermore, though the recording of ethnicity on death certificates has been the norm in Scotland for the past decade, it is only now being considered for use in England.^42–44^

## Conclusions

Ethnic minority groups have suffered the consequences of the COVID-19 epidemic, disproportionately, in the UK. Economic deprivation and underlying health conditions do not fully explain this adverse effect on major groups within English Society. We need better, and more readily available, linked data to be able to characterise ethnic disparities in more detail, and investigate in detail whether discrimination, access to protective equipment, lifestyle, behavioural factors, or access to health care are important factors. Engaging with ethnic minority communities to understand their lived experiences will be essential for generating evidence to prevent further widening of inequalities in a timely and actionable manner. Equality is difficult to achieve, but structural and persistent inequalities must be addressed in a civilised society.

## Data Availability

NHS England is the data controller; TPP is the data processor; and the key researchers on OpenSAFELY are acting on behalf of NHS England. This implementation of OpenSAFELY is hosted within the TPP environment which is accredited to the ISO 27001 information security standard and is NHS IG Toolkit compliant; patient data has been pseudonymised for analysis and linkage using industry standard cryptographic hashing techniques; all pseudonymised datasets transmitted for linkage onto OpenSAFELY are encrypted; access to the platform is via a virtual private network (VPN) connection, restricted to a small group of researchers; the researchers hold contracts with NHS England and only access the platform to initiate database queries and statistical models; all database activity is logged; only aggregate statistical outputs leave the platform environment following best practice for anonymisation of results such as statistical disclosure control for low cell counts. Detailed information on all code lists is openly shared at https://codelists.opensafely.org/ for inspection and reuse.

https://github.com/opensafely/ethnicity-covid-research

https://codelists.opensafely.org/

## Acknowledgements

We are very grateful for all the support received from the TPP Technical Operations team throughout this work, and for generous assistance from the information governance and database teams at NHS England / NHSX.

## Information governance and ethical approval

NHS England is the data controller; TPP is the data processor; and the key researchers on OpenSAFELY are acting on behalf of NHS England. This implementation of OpenSAFELY is hosted within the TPP environment which is accredited to the ISO 27001 information security standard and is NHS IG Toolkit compliant; patient data has been pseudonymised for analysis and linkage using industry standard cryptographic hashing techniques; all pseudonymised datasets transmitted for linkage onto OpenSAFELY are encrypted; access to the platform is via a virtual private network (VPN) connection, restricted to a small group of researchers; the researchers hold contracts with NHS England and only access the platform to initiate database queries and statistical models; all database activity is logged; only aggregate statistical outputs leave the platform environment following best practice for anonymisation of results such as statistical disclosure control for low cell counts. The OpenSAFELY research platform adheres to the data protection principles of the UK Data Protection Act 2018 and the EU General Data Protection Regulation (GDPR) 2016. In March 2020, the Secretary of State for Health and Social Care used powers under the UK Health Service (Control of Patient Information) Regulations 2002 (COPI) to require organisations to process confidential patient information for the purposes of protecting public health, providing healthcare services to the public and monitoring and managing the COVID-19 outbreak and incidents of exposure.[4] Taken together, these provide the legal bases to link patient datasets on the OpenSAFELY platform. GP practices, from which the primary care data are obtained, are required to share relevant health information to support the public health response to the pandemic and have been informed of the OpenSAFELY analytics platform.

This study was approved by the Health Research Authority (REC reference 20/LO/0651) and by the LSHTM Ethics Board (reference 21863).

## Guarantor

RM/LS/BG are guarantors

## Contributorship

Contributions are as follows:

Conceptualization: RM, CTR, KB, RME, LS, BG, BM, HJC, SJWE, KK, DH, KR

Data curation: RM, CTR, AJW, CB, JC, CM, RME, WJH, BM, SB

Formal analysis: RM, CTR

Funding acquisition: LS, BG, RME

Investigation: RM, CTR, CM, WJH

Methodology: RM, CTR, KB, RME, KK, NC, RG, DH, KR, LS, BG, BM, EW, HJC, SJWE

Codelists: RM, LT, AS, AJW, CM, BG, WJH, SB, AM

Project administration: RM, CTR, AS, AJW, CM, BG, WJH

Resources: CB JC BG BM SB AM

Software: AJW CB JC DE PI CM WJH BN SB HJC ND RC JP FH SH

Visualisation: RM RME

Writing - original draft: RM

Writing- review & editing: ALL

Information governance: CB LS BG AM

## Supplementary Appendix

### Open source materials

Open source materials: All code for data management and analyses and raw outputs are openly shared online for review and re-use (https://github.com/opensafely/ethnicity-covid-research). All iterations of the pre-specified study protocol are archived with version control (https://github.com/opensafely/ethnicity-covid-research/tree/master/protocol).

### Information governance and ethics

NHS England is the data controller; TPP is the data processor; and the key researchers on OpenSAFELY are acting on behalf of NHS England. OpenSAFELY is hosted within the TPP environment which is accredited to the ISO 27001 information security standard and is NHS IG Toolkit compliant;1,2 patient data are pseudonymised for analysis and linkage using industry standard cryptographic hashing techniques; all pseudonymised datasets transmitted for linkage onto OpenSAFELY are encrypted; access to the platform is via a virtual private network (VPN) connection, restricted to a small group of researchers who hold contracts with NHS England and only access the platform to initiate database queries and statistical models. The platform includes pseudonymized data such as coded diagnoses, medications and physiological parameters. No free text data are included. This, in addition to other technical and organisational controls, minimizes any risk of re-identification. Similarly pseudonymized datasets from other data providers are securely provided to the EHR vendor and linked to the primary care data.

All database activity is logged; only aggregate statistical outputs leave the platform environment following best practice for anonymization of results such as statistical disclosure control for low cell counts.3 The OpenSAFELY platform adheres to the data protection principles of the UK Data Protection Act 2018 and the EU General Data Protection Regulation (GDPR) 2016. In March 2020, the Secretary of State for Health and Social Care used powers under the UK Health Service (Control of Patient Information) Regulations 2002 (COPI) to require organisations to process confidential patient information for the purposes of protecting public health, providing healthcare services to the public and monitoring and managing the COVID-19 outbreak and incidents of exposure.4 Taken together, these provide the legal bases to link patient datasets on the OpenSAFELY platform. This study was approved by the Health Research Authority (REC reference 20/LO/0651) and by the LSHTM Ethics Board (ref 21863).

### Patient and public involvement

Patients were not formally involved in developing this specific study design that was developed rapidly in the context of a global health emergency. We have developed a publicly available website https://opensafely.org/ through which we invite any patient or member of the public to contact us regarding this study or the broader OpenSAFELY project.

**Table S1.**
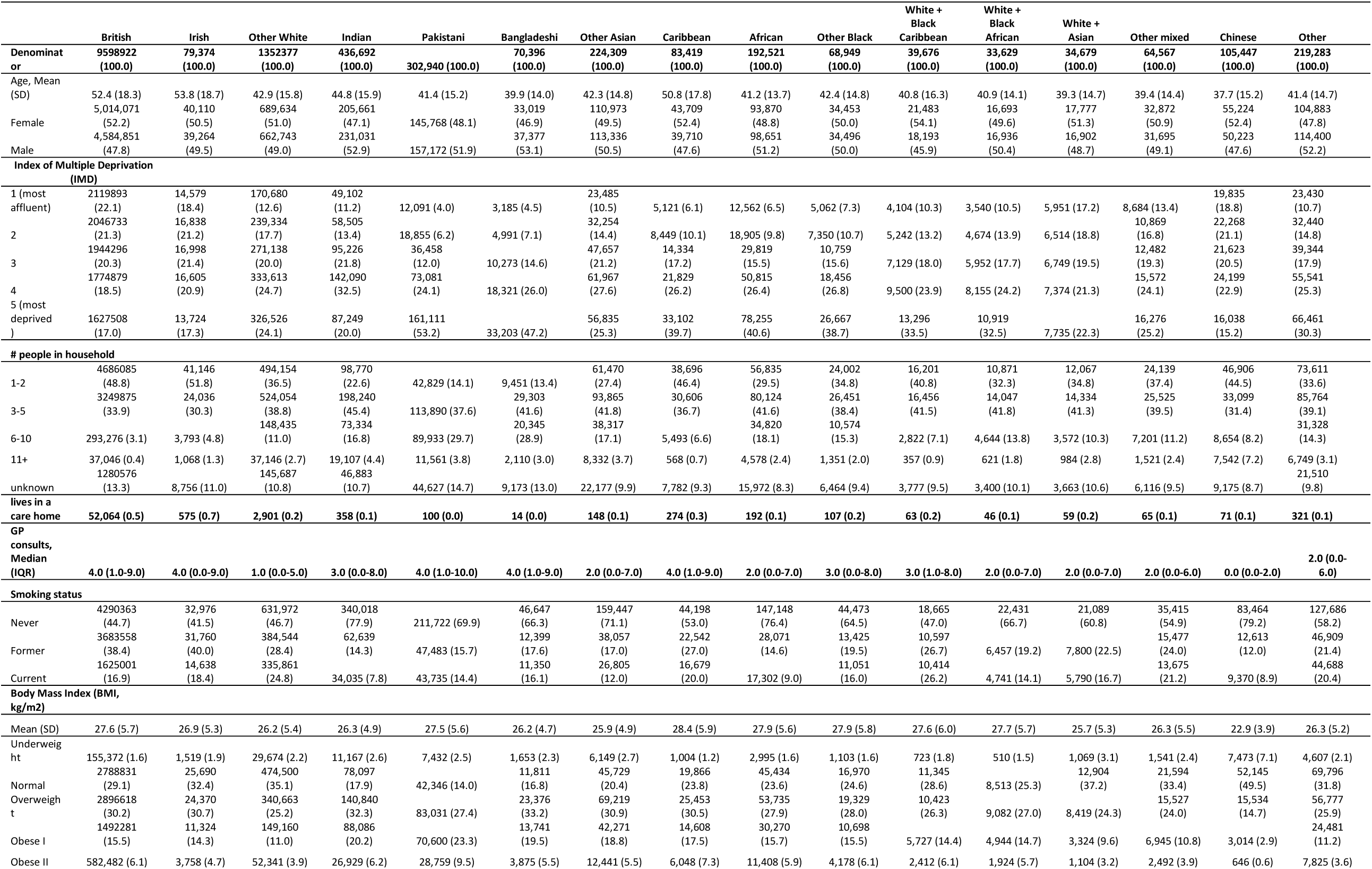
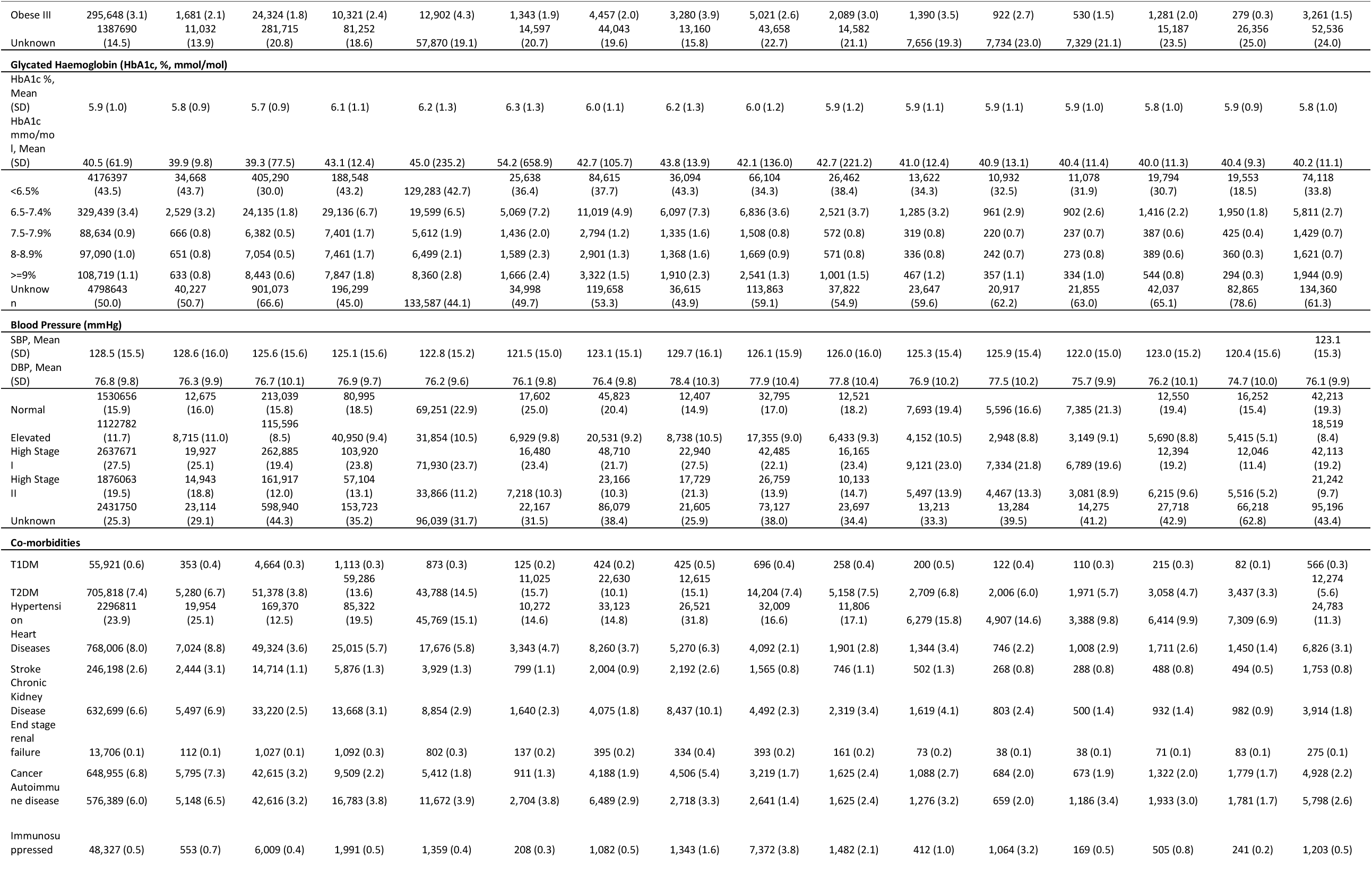
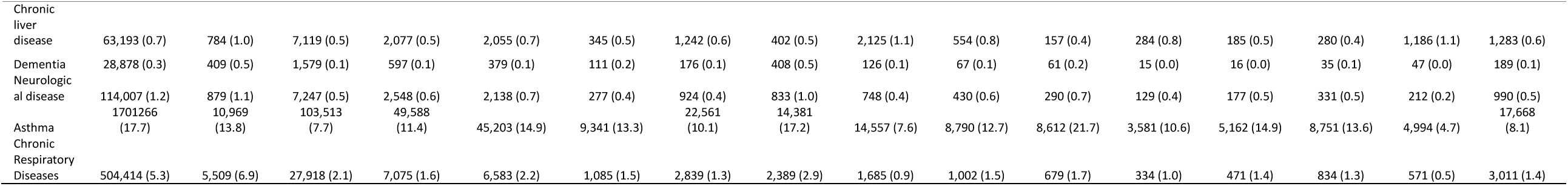
Baseline characteristics by ethnic group in 16 categories.

**Table S2.**
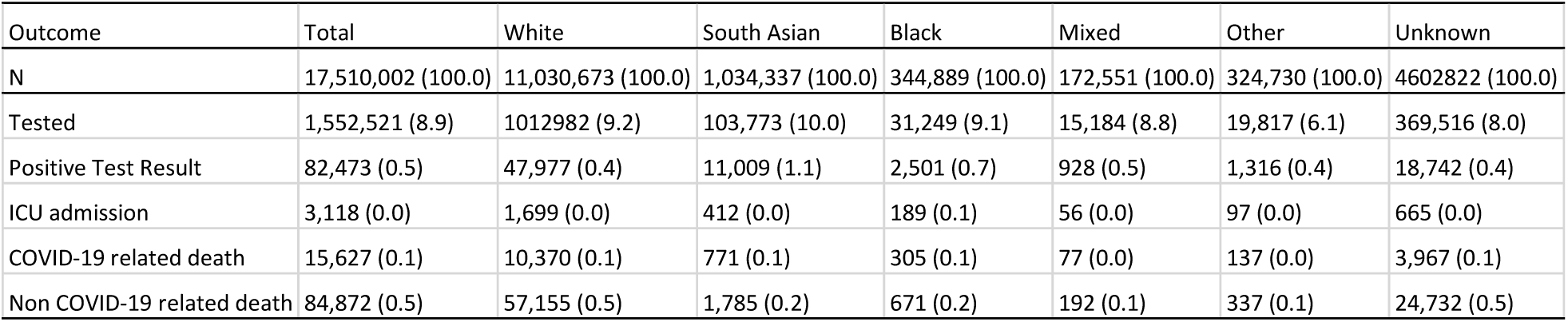
Count of outcomes by ethnic group in five categories.

| Outcome | Total | White | South Asian | Black | Mixed | Other | Unknown |
| --- | --- | --- | --- | --- | --- | --- | --- |
| N | 17,510,002 (100.0) | 11,030,673 (100.0) | 1,034,337 (100.0) | 344,889 (100.0) | 172,551 (100.0) | 324,730 (100.0) | 4602822 (100.0) |
| Tested | 1,552,521 (8.9) | 1012982 (9.2) | 103,773 (10.0) | 31,249 (9.1) | 15,184 (8.8) | 19,817 (6.1) | 369,516 (8.0) |
| Positive Test Result | 82,473 (0.5) | 47,977 (0.4) | 11,009 (1.1) | 2,501 (0.7) | 928 (0.5) | 1,316 (0.4) | 18,742 (0.4) |
| ICU admission | 3,118 (0.0) | 1,699 (0.0) | 412 (0.0) | 189 (0.1) | 56 (0.0) | 97 (0.0) | 665 (0.0) |
| COVID-19 related death | 15,627 (0.1) | 10,370 (0.1) | 771 (0.1) | 305 (0.1) | 77 (0.0) | 137 (0.0) | 3,967 (0.1) |
| Non COVID-19 related death | 84,872 (0.5) | 57,155 (0.5) | 1,785 (0.2) | 671 (0.2) | 192 (0.1) | 337 (0.1) | 24,732 (0.5) |

**Table S3.** Count of outcomes by ethnic group in 16 categories.

|  | British | Irish | Other White | White +<br>Black<br>Caribbean | White +<br>Black African | White +<br>Asian | Other mixed | Indian | Pakistani | Bangladeshi | Other Asian | Caribbean | African |
| --- | --- | --- | --- | --- | --- | --- | --- | --- | --- | --- | --- | --- | --- |
| N | 9,598,922<br>(100.0) | 79,374<br>(100.0) | 1,352,377<br>(100.0) | 436,692<br>(100.0) | 302,940<br>(100.0) | 70,396<br>(100.0) | 224,309<br>(100.0) | 83,419<br>(100.0) | 192,521<br>(100.0) | 68,949<br>(100.0) | 105,447<br>(100.0) | 172,551<br>(100.0) | 219,283<br>(100.0) |
| Tested | 911,140<br>(9.5) | 7,929 (10.0) | 93,913 (6.9) | 50,068<br>(11.5) | 28,586 (9.4) | 4,951 (7.0) | 20,168 (9.0) | 7,442 (8.9) | 17,928 (9.3) | 5,879 (8.5) | 3,663 (3.5) | 15,184 (8.8) | 16,154 (7.4) |
| Positive Test<br>Result | 42,328 (0.4) | 412 (0.5) | 5,237 (0.4) | 4,539 (1.0) | 4,088 (1.3) | 426 (0.6) | 1,956 (0.9) | 590 (0.7) | 1,518 (0.8) | 393 (0.6) | 151 (0.1) | 928 (0.5) | 1,165 (0.5) |
| ICU admission | [REDACTED] |  |  |  |  |  |  |  |  |  |  |  |  |
| COVID-19 related<br>death | 9,683 (0.1) | 127 (0.2) | 560 (0.0) | 357 (0.1) | 214 (0.1) | 49 (0.1) | 151 (0.1) | 164 (0.2) | 100 (0.1) | 41 (0.1) | 28 (0.0) | 77 (0.0) | 109 (0.0) |
| Non COVID-19<br>related death | 53,687 (0.6) | 554 (0.7) | 2,914 (0.2) | 891 (0.2) | 529 (0.2) | 93 (0.1) | 272 (0.1) | 389 (0.5) | 174 (0.1) | 108 (0.2) | 66 (0.1) | 192 (0.1) | 271 (0.1) |

**Table S4.** Association between ethnicity in 5 categories and COVID-19 outcomes (serial adjustment)

|  | Denominator | Event | Total person-weeks | Rate per 1,000 | Crude |  | Age-sex adjusted |  | plus IMD |  | plus co-morbidities, GP consults |  | plus household size and care home status |  |
| --- | --- | --- | --- | --- | --- | --- | --- | --- | --- | --- | --- | --- | --- | --- |
|  |  |  |  |  | HR | 95% CI | HR | 95% CI | HR | 95% CI | HR | 95% CI | HR | 95% CI |
| **Tested** |  |  |  |  |  |  |  |  |  |  |  |  |  |  |
| White | 11,030,673 | 834,665 | 283,175,442 | 2.95 | 1 |  | 1 |  | 1 |  | 1 |  | 1 |  |
| South Asian | 1,034,337 | 83,876 | 26,603,024 | 3.15 | 1.05 | (1.05 - 1.06) | 1.1 | (1.09 - 1.11) | 1.07 | (1.06 - 1.08) | 1.07 | (1.06 - 1.08) | 1.08 | (1.07 - 1.09) |
| Black | 344,889 | 25,909 | 8,859,029 | 2.92 | 1.02 | (1.01 - 1.03) | 1.07 | (1.06 - 1.09) | 1.04 | (1.02 - 1.05) | 1.07 | (1.06 - 1.08) | 1.08 | (1.06 - 1.09) |
| Mixed | 172,551 | 12,418 | 4,443,402 | 2.79 | 0.97 | (0.95 - 0.99) | 1.03 | (1.01 - 1.05) | 1.01 | (0.99 - 1.03) | 1.03 | (1.01 - 1.05) | 1.03 | (1.01 - 1.05) |
| Other | 324,730 | 16,256 | 8,411,306 | 1.93 | 0.67 | (0.66 - 0.68) | 0.71 | (0.70 - 0.72) | 0.70 | (0.69 - 0.71) | 0.76 | (0.75 - 0.77) | 0.76 | (0.74 - 0.77) |
| Unknown | 4,602,822 | 302,545 | 118,472,512 | 2.55 | 0.86 | (0.86 - 0.87) | 0.88 | (0.88 - 0.89) | 0.88 | (0.88 - 0.89) | 0.96 | (0.95 - 0.96) | 0.96 | (0.96 - 0.97) |
| **Positive Test** |  |  |  |  |  |  |  |  |  |  |  |  |  |  |
| White | 11,030,673 | 46,136 | 288,476,946 | 0.16 | 1 |  | 1 |  | 1 |  | 1 |  | 1 |  |
| South Asian | 1,034,337 | 9,815 | 27,059,833 | 0.36 | 2.18 | (2.13 - 2.23) | 2.53 | (2.47 - 2.59) | 2.35 | (2.29 - 2.41) | 2.11 | (2.06 - 2.16) | 2.02 | (1.97 - 2.07) |
| Black | 344,889 | 2,358 | 9,022,706 | 0.26 | 1.69 | (1.62 - 1.76) | 1.98 | (1.90 - 2.07) | 1.8 | (1.72 - 1.88) | 1.69 | (1.62 - 1.77) | 1.68 | (1.61 - 1.76) |
| Mixed | 172,551 | 865 | 4,521,307 | 0.19 | 1.26 | (1.18 - 1.35) | 1.56 | (1.46 - 1.67) | 1.48 | (1.39 - 1.59) | 1.47 | (1.37 - 1.57) | 1.46 | (1.36 - 1.56) |
| Other | 324,730 | 1,257 | 8,513,352 | 0.15 | 0.98 | (0.93 - 1.04) | 1.2 | (1.14 - 1.27) | 1.16 | (1.09 - 1.23) | 1.19 | (1.13 - 1.26) | 1.16 | (1.10 - 1.23) |
| Unknown | 4,602,822 | 17,783 | 120,384,887 | 0.15 | 0.96 | (0.94 - 0.98) | 0.98 | (0.96 - 0.99) | 0.98 | (0.97 - 1.00) | 1.02 | (1.00 - 1.04) | 1.03 | (1.01 - 1.05) |
| **ICU admission** |  |  |  |  |  |  |  |  |  |  |  |  |  |  |
| White | 11,030,673 | 1,697 | 282,702,976 | 0.01 | 1 |  | 1 |  | 1 |  | 1 |  | 1 |  |
| South Asian | 1,034,337 | 410 | 26,558,295 | 0.02 | 2.38 | (2.12 - 2.66) | 3.29 | (2.93 - 3.70) | 3.05 | (2.71 - 3.43) | 2.38 | (2.11 - 2.69) | 2.22 | (1.96 - 2.52) |
| Black | 344,889 | 188 | 8,853,078 | 0.02 | 3.09 | (2.65 - 3.61) | 3.93 | (3.36 - 4.60) | 3.51 | (2.99 - 4.12) | 3.16 | (2.68 - 3.71) | 3.07 | (2.61 - 3.61) |
| Mixed | 172,551 | 56 | 4,432,680 | 0.01 | 1.96 | (1.50 - 2.56) | 3.17 | (2.42 - 4.14) | 3.00 | (2.29 - 3.93) | 2.92 | (2.23 - 3.83) | 2.86 | (2.19 - 3.75) |
| Other | 324,730 | 97 | 8,342,471 | 0.01 | 1.82 | (1.48 - 2.24) | 2.9 | (2.35 - 3.58) | 2.79 | (2.26 - 3.44) | 2.95 | (2.39 - 3.64) | 2.86 | (2.31 - 3.53) |
| Unknown | 4,602,822 | 665 | 117,954,816 | 0.01 | 0.95 | (0.87 - 1.04) | 1.03 | (0.94 - 1.12) | 1.03 | (0.94 - 1.13) | 0.97 | (0.89 - 1.07) | 0.97 | (0.88 - 1.07) |
| **COVID-19 related death** |  |  |  |  |  |  |  |  |  |  |  |  |  |  |
| White | 11,030,673 | 10,370 | 288,983,863 | 0.04 | 1 |  | 1 |  | 1 |  | 1 |  | 1 |  |
| South Asian | 1,034,337 | 771 | 27,151,760 | 0.03 | 0.71 | (0.66 - 0.77) | 1.58 | (1.46 - 1.70) | 1.39 | (1.29 - 1.51) | 1.28 | (1.19 - 1.39) | 1.27 | (1.17 - 1.38) |
| Black | 344,889 | 305 | 9,051,561 | 0.03 | 0.84 | (0.75 - 0.94) | 1.72 | (1.53 - 1.94) | 1.47 | (1.31 - 1.66) | 1.37 | (1.22 - 1.54) | 1.55 | (1.38 - 1.75) |
| Mixed | 172,551 | 77 | 4,531,702 | 0.02 | 0.45 | (0.36 - 0.56) | 1.49 | (1.19 - 1.87) | 1.37 | (1.10 - 1.72) | 1.37 | (1.09 - 1.72) | 1.40 | (1.12 - 1.76) |
| Other | 324,730 | 137 | 8,528,791 | 0.02 | 0.43 | (0.36 - 0.51) | 1.28 | (1.08 - 1.52) | 1.20 | (1.01 - 1.43) | 1.23 | (1.04 - 1.46) | 1.25 | (1.05 - 1.49) |
| Unknown | 4,602,822 | 3,967 | 120,574,720 | 0.03 | 0.93 | (0.90 - 0.96) | 0.88 | (0.85 - 0.91) | 0.89 | (0.86 - 0.92) | 0.96 | (0.92 - 1.00) | 1.01 | (0.97 - 1.05) |

| Non-COVID related death |  |  |  |  |  |  |  |  |  |  |  |  |  |  |
| --- | --- | --- | --- | --- | --- | --- | --- | --- | --- | --- | --- | --- | --- | --- |
| White | 11,030,673 | 57,155 | 288,983,863 | 0.2 | 1 |  | 1 |  | 1 |  | 1 |  | 1 |  |
| South Asian | 1,034,337 | 1,785 | 27,151,760 | 0.07 | 0.37 | (0.35 - 0.39) | 0.81 | (0.78 - 0.86) | 0.74 | (0.70 - 0.77) | 0.82 | (0.78 - 0.86) | 0.82 | (0.78 - 0.87) |
| Black | 344,889 | 671 | 9,051,561 | 0.07 | 0.42 | (0.39 - 0.46) | 0.89 | (0.82 - 0.96) | 0.78 | (0.72 - 0.84) | 0.83 | (0.77 - 0.90) | 0.87 | (0.80 - 0.94) |
| Mixed | 172,551 | 192 | 4,531,702 | 0.04 | 0.23 | (0.20 - 0.27) | 0.77 | (0.67 - 0.89) | 0.72 | (0.62 - 0.83) | 0.78 | (0.68 - 0.90) | 0.79 | (0.69 - 0.91) |
| Other | 324,730 | 337 | 8,528,791 | 0.04 | 0.22 | (0.20 - 0.25) | 0.67 | (0.60 - 0.74) | 0.64 | (0.58 - 0.71) | 0.71 | (0.64 - 0.80) | 0.72 | (0.64 - 0.80) |
| Unknown | 4,602,822 | 24,732 | 120,574,720 | 0.21 | 1.03 | (1.02 - 1.05) | 0.96 | (0.95 - 0.98) | 0.97 | (0.96 - 0.99) | 1.07 | (1.05 - 1.09) | 1.10 | (1.08 - 1.12) |

**Table S5.** Association between ethnicity in 16 categories and COVID-19 outcomes (serial adjustment)

|  | Denominator | Event | Total person-weeks | Rate per 1,000 | Crude |  | Age-sex adjusted |  | plus IMD |  | plus co-morbidities, GP consults |  | plus household size and carehome |  |
| --- | --- | --- | --- | --- | --- | --- | --- | --- | --- | --- | --- | --- | --- | --- |
|  |  |  |  |  | HR | 95% CI | HR | 95% CI | HR | 95% CI | HR | 95% CI | HR | 95% CI |
| **Tested** |  |  |  |  |  |  |  |  |  |  |  |  |  |  |
| British | 9,598,922 | 751,934 | 246,165,394 | 3.05 | 1 |  | 1 |  | 1 |  | 1 |  | 1 |  |
| Irish | 79,374 | 6,537 | 2,030,922 | 3.22 | 1.09 | (1.07 - 1.12) | 1.06 | (1.03 - 1.09) | 1.05 | (1.03 - 1.08) | 1.05 | (1.02 - 1.08) | 1.05 | (1.02 - 1.07) |
| Other White | 1,352,377 | 76,194 | 34,979,125 | 2.18 | 0.73 | (0.72 - 0.73) | 0.74 | (0.74 - 0.75) | 0.73 | (0.72 - 0.74) | 0.79 | (0.79 - 0.80) | 0.79 | (0.79 - 0.80) |
| White + Black Caribbean | 39,676 | 2,912 | 4,443,402 | 0.66 | 0.95 | (0.91 - 0.98) | 0.99 | (0.95 - 1.02) | 0.96 | (0.93 - 1.00) | 0.95 | (0.92 - 0.99) | 0.96 | (0.92 - 0.99) |
| White + Black African | 33,629 | 2,676 | 4,443,402 | 0.6 | 1.03 | (0.99 - 1.07) | 1.1 | (1.06 - 1.14) | 1.07 | (1.03 - 1.11) | 1.14 | (1.10 - 1.18) | 1.13 | (1.09 - 1.17) |
| White + Asian | 34,679 | 2,674 | 4,443,402 | 0.6 | 0.99 | (0.95 - 1.03) | 1.05 | (1.01 - 1.09) | 1.04 | (1.00 - 1.08) | 1.06 | (1.02 - 1.11) | 1.06 | (1.02 - 1.10) |
| Other mixed | 64,567 | 4,156 | 4,443,402 | 0.94 | 0.84 | (0.81 - 0.87) | 0.88 | (0.86 - 0.91) | 0.87 | (0.84 - 0.89) | 0.9 | (0.87 - 0.93) | 0.9 | (0.87 - 0.93) |
| Indian | 436,692 | 40,924 | 11,204,952 | 3.65 | 1.1 | (1.09 - 1.11) | 1.14 | (1.13 - 1.15) | 1.11 | (1.10 - 1.12) | 1.15 | (1.14 - 1.16) | 1.15 | (1.13 - 1.16) |
| Pakistani | 302,940 | 22,162 | 7,815,500 | 2.84 | 0.96 | (0.95 - 0.97) | 1 | (0.99 - 1.02) | 0.96 | (0.95 - 0.97) | 0.94 | (0.93 - 0.96) | 0.95 | (0.94 - 0.96) |
| Bangladeshi | 70,396 | 3,962 | 1,823,245 | 2.17 | 0.7 | (0.68 - 0.72) | 0.74 | (0.72 - 0.76) | 0.71 | (0.68 - 0.73) | 0.69 | (0.67 - 0.71) | 0.7 | (0.67 - 0.72) |
| Other Asian | 224,309 | 16,828 | 5,759,327 | 2.92 | 0.98 | (0.97 - 1.00) | 1.03 | (1.01 - 1.05) | 1 | (0.99 - 1.02) | 1.05 | (1.03 - 1.07) | 1.05 | (1.03 - 1.07) |
| Caribbean | 83,419 | 6,191 | 2,139,363 | 2.89 | 0.99 | (0.97 - 1.02) | 0.99 | (0.97 - 1.02) | 0.96 | (0.93 - 0.98) | 0.96 | (0.94 - 0.99) | 0.98 | (0.95 - 1.00) |
| African | 192,521 | 14,899 | 4,944,673 | 3.01 | 0.99 | (0.98 - 1.01) | 1.06 | (1.04 - 1.08) | 1.01 | (1.00 - 1.03) | 1.09 | (1.07 - 1.11) | 1.09 | (1.07 - 1.10) |
| Other Black | 68,949 | 4,819 | 1,774,993 | 2.71 | 0.91 | (0.88 - 0.93) | 0.97 | (0.94 - 1.00) | 0.94 | (0.91 - 0.96) | 0.96 | (0.94 - 0.99) | 0.96 | (0.94 - 0.99) |
| Chinese | 105,447 | 2,995 | 2,748,718 | 1.09 | 0.35 | (0.34 - 0.36) | 0.37 | (0.36 - 0.38) | 0.37 | (0.36 - 0.38) | 0.44 | (0.42 - 0.45) | 0.43 | (0.42 - 0.45) |
| Other | 219,283 | 13,261 | 5,662,588 | 2.34 | 0.79 | (0.77 - 0.80) | 0.83 | (0.81 - 0.84) | 0.81 | (0.79 - 0.82) | 0.86 | (0.85 - 0.88) | 0.86 | (0.84 - 0.87) |
| Unknown | 4,602,822 | 302,545 | 118,472,512 | 2.55 | 0.84 | (0.83 - 0.84) | 0.86 | (0.85 - 0.86) | 0.86 | (0.85 - 0.86) | 0.93 | (0.93 - 0.94) | 0.94 | (0.94 - 0.95) |
| **Positive Test** |  |  |  |  |  |  |  |  |  |  |  |  |  |  |
| British | 9,598,922 | 40,845 | 250,960,238 | 0.16 | 1 |  | 1 |  | 1 |  | 1 |  | 1 |  |
| Irish | 79,374 | 398 | 2,072,202 | 0.19 | 1.3 | (1.18 - 1.44) | 1.21 | (1.09 - 1.33) | 1.18 | (1.07 - 1.30) | 1.18 | (1.06 - 1.30) | 1.18 | (1.07 - 1.30) |
| Other White | 1,352,377 | 4,893 | 35,444,512 | 0.14 | 0.93 | (0.90 - 0.96) | 1.08 | (1.04 - 1.11) | 1.02 | (0.99 - 1.06) | 1.11 | (1.07 - 1.14) | 1.1 | (1.06 - 1.13) |
| White + Black Caribbean | 39,676 | 177 | 4,521,307 | 0.04 | 1.08 | (0.93 - 1.25) | 1.31 | (1.13 - 1.52) | 1.23 | (1.06 - 1.43) | 1.22 | (1.05 - 1.41) | 1.24 | (1.07 - 1.44) |
| White + Black African | 33,629 | 228 | 4,521,307 | 0.05 | 1.66 | (1.46 - 1.89) | 2.11 | (1.86 - 2.41) | 1.95 | (1.71 - 2.23) | 1.94 | (1.70 - 2.21) | 1.89 | (1.66 - 2.16) |
| White + Asian | 34,679 | 200 | 4,521,307 | 0.04 | 1.43 | (1.25 - 1.64) | 1.83 | (1.59 - 2.10) | 1.78 | (1.55 - 2.05) | 1.76 | (1.53 - 2.03) | 1.71 | (1.49 - 1.97) |
| Other mixed | 64,567 | 260 | 4,521,307 | 0.06 | 1.04 | (0.92 - 1.18) | 1.32 | (1.17 - 1.50) | 1.26 | (1.11 - 1.42) | 1.29 | (1.14 - 1.45) | 1.27 | (1.13 - 1.44) |
| Indian | 436,692 | 4,111 | 11,421,323 | 0.36 | 2.16 | (2.08 - 2.23) | 2.48 | (2.39 - 2.56) | 2.34 | (2.26 - 2.42) | 2.16 | (2.08 - 2.23) | 2.07 | (2.00 - 2.15) |
| Pakistani | 302,940 | 3,487 | 7,925,011 | 0.44 | 2.4 | (2.32 - 2.49) | 2.89 | (2.79 - 2.99) | 2.58 | (2.48 - 2.67) | 2.27 | (2.18 - 2.35) | 2.14 | (2.06 - 2.22) |
| Bangladeshi | 70,396 | 369 | 1,844,952 | 0.2 | 1.2 | (1.09 - 1.33) | 1.49 | (1.35 - 1.66) | 1.32 | (1.19 - 1.47) | 1.16 | (1.04 - 1.29) | 1.11 | (1.00 - 1.23) |
| Other Asian | 224,309 | 1,848 | 5,868,546 | 0.31 | 2.11 | (2.01 - 2.21) | 2.58 | (2.46 - 2.70) | 2.42 | (2.31 - 2.54) | 2.31 | (2.20 - 2.42) | 2.23 | (2.13 - 2.34) |
| Caribbean | 83,419 | 568 | 2,178,227 | 0.26 | 1.7 | (1.57 - 1.85) | 1.72 | (1.59 - 1.88) | 1.56 | (1.43 - 1.69) | 1.46 | (1.34 - 1.59) | 1.53 | (1.40 - 1.66) |
| African | 192,521 | 1,416 | 5,038,797 | 0.28 | 1.77 | (1.68 - 1.87) | 2.26 | (2.14 - 2.39) | 2.03 | (1.93 - 2.15) | 1.95 | (1.84 - 2.06) | 1.9 | (1.79 - 2.00) |
| Other Black | 68,949 | 374 | 1,805,682 | 0.21 | 1.34 | (1.21 - 1.49) | 1.69 | (1.53 - 1.88) | 1.54 | (1.39 - 1.71) | 1.49 | (1.34 - 1.65) | 1.45 | (1.31 - 1.61) |
| Chinese | 105,447 | 146 | 2,768,474 | 0.05 | 0.34 | (0.29 - 0.40) | 0.44 | (0.37 - 0.52) | 0.44 | (0.38 - 0.52) | 0.48 | (0.41 - 0.57) | 0.47 | (0.40 - 0.55) |
| Other | 219,283 | 1,111 | 5,744,878 | 0.19 | 1.29 | (1.21 - 1.37) | 1.58 | (1.49 - 1.68) | 1.48 | (1.40 - 1.58) | 1.52 | (1.43 - 1.62) | 1.48 | (1.40 - 1.58) |
| Unknown | 4,602,822 | 17,783 | 120,384,887 | 0.15 | 0.95 | (0.94 - 0.97) | 0.99 | (0.97 - 1.00) | 0.99 | (0.97 - 1.00) | 1.03 | (1.01 - 1.05) | 1.04 | (1.02 - 1.06) |
| **ICU admission** |  |  |  |  |  |  |  |  |  |  |  |  |  |  |
| British | 9,598,922 | 1,507 | 245,945,765 | 0.01 | 1 |  | 1 |  | 1 |  | 1 |  | 1 |  |
| Irish | 79,374 | . | 2,031,511 | . | 0.36 | (0.15 - 0.88) | 0.36 | (0.15 - 0.88) | 0.35 | (0.15 - 0.85) | 0.37 | (0.15 - 0.89) | 0.37 | (0.15 - 0.89) |
| Other White | 1,352,377 | 185 | 34,725,703 | 0.01 | 0.83 | (0.71 - 0.96) | 1.3 | (1.11 - 1.52) | 1.25 | (1.07 - 1.46) | 1.4 | (1.20 - 1.64) | 1.38 | (1.18 - 1.61) |
| White + Black Caribbean | 39,676 | 11 | 4,432,680 | 0 | 1.6 | (0.88 - 2.89) | 2.61 | (1.44 - 4.73) | 2.42 | (1.34 - 4.39) | 2.36 | (1.30 - 4.28) | 2.34 | (1.29 - 4.25) |
| White + Black African | 33,629 | 21 | 4,432,680 | 0 | 3.58 | (2.32 - 5.50) | 5.65 | (3.67 - 8.71) | 5.21 | (3.38 - 8.03) | 5 | (3.24 - 7.71) | 4.84 | (3.14 - 7.47) |
| White + Asian | 34,679 | 10 | 4,432,680 | 0 | 1.73 | (0.93 - 3.23) | 3.15 | (1.69 - 5.87) | 3.08 | (1.65 - 5.74) | 3.11 | (1.67 - 5.80) | 3.03 | (1.63 - 5.66) |
| Other mixed | 64,567 | 14 | 4,432,680 | 0 | 1.26 | (0.74 - 2.14) | 2.32 | (1.37 - 3.93) | 2.22 | (1.31 - 3.76) | 2.25 | (1.33 - 3.82) | 2.2 | (1.30 - 3.73) |
| Indian | 436,692 | 161 | 11,210,534 | 0.01 | 2.18 | (1.84 - 2.59) | 2.91 | (2.45 - 3.47) | 2.77 | (2.33 - 3.29) | 2.23 | (1.86 - 2.66) | 2.1 | (1.75 - 2.51) |
| Pakistani | 302,940 | 118 | 7,778,925 | 0.02 | 2.22 | (1.83 - 2.69) | 3.55 | (2.92 - 4.31) | 3.12 | (2.56 - 3.80) | 2.33 | (1.91 - 2.84) | 2.14 | (1.74 - 2.62) |
| Bangladeshi | 70,396 | 25 | 1,807,946 | 0.01 | 1.96 | (1.31 - 2.91) | 3.49 | (2.34 - 5.21) | 3.07 | (2.06 - 4.58) | 2.33 | (1.56 - 3.48) | 2.15 | (1.44 - 3.22) |
| Other Asian | 224,309 | 106 | 5,760,890 | 0.02 | 2.7 | (2.20 - 3.30) | 4.16 | (3.39 - 5.10) | 3.92 | (3.19 - 4.81) | 3.37 | (2.74 - 4.14) | 3.2 | (2.60 - 3.95) |
| Caribbean | 83,419 | 42 | 2,137,043 | 0.02 | 2.72 | (1.99 - 3.71) | 2.82 | (2.06 - 3.85) | 2.49 | (1.82 - 3.42) | 2.28 | (1.66 - 3.12) | 2.26 | (1.65 - 3.10) |
| African | 192,521 | 118 | 4,945,421 | 0.02 | 3.37 | (2.79 - 4.08) | 5.21 | (4.29 - 6.32) | 4.67 | (3.84 - 5.68) | 4.25 | (3.48 - 5.18) | 4.07 | (3.33 - 4.97) |
| Other Black | 68,949 | 28 | 1,770,614 | 0.02 | 2.22 | (1.52 - 3.23) | 3.14 | (2.15 - 4.57) | 2.75 | (1.88 - 4.04) | 2.58 | (1.76 - 3.79) | 2.51 | (1.71 - 3.69) |
| Chinese | 105,447 | 18 | 2,709,895 | 0.01 | 1.01 | (0.63 - 1.60) | 2.01 | (1.26 - 3.21) | 2.02 | (1.26 - 3.21) | 2.51 | (1.57 - 4.00) | 2.44 | (1.53 - 3.89) |
| Other | 219,283 | 79 | 5,632,575 | 0.01 | 2.11 | (1.68 - 2.66) | 3.38 | (2.68 - 4.26) | 3.18 | (2.52 - 4.02) | 3.27 | (2.59 - 4.13) | 3.17 | (2.51 - 4.01) |
| Unknown | 4,602,822 | 665 | 117,954,816 | 0.01 | 0.92 | (0.84 - 1.01) | 1.05 | (0.96 - 1.15) | 1.05 | (0.95 - 1.15) | 1 | (0.91 - 1.10) | 1 | (0.91 - 1.10) |
| **COVID-19 related death** |  |  |  |  |  |  |  |  |  |  |  |  |  |  |
| British | 9,598,922 | 9,683 | 251,408,914 | 0.04 | 1 |  | 1 |  | 1 |  | 1 |  | 1 |  |
| Irish | 79,374 | 127 | 2,076,507 | 0.06 | 1.45 | (1.22 - 1.73) | 1.14 | (0.95 - 1.36) | 1.07 | (0.90 - 1.27) | 1.01 | (0.85 - 1.21) | 1.02 | (0.85 - 1.21) |
| Other White | 1,352,377 | 560 | 35,498,448 | 0.02 | 0.39 | (0.36 - 0.43) | 0.89 | (0.81 - 0.97) | 0.86 | (0.79 - 0.93) | 0.89 | (0.81 - 0.97) | 0.92 | (0.85 - 1.01) |
| White + Black Caribbean | 39,676 | 31 | 4,531,702 | 0.01 | 0.69 | (0.48 - 0.98) | 1.74 | (1.22 - 2.48) | 1.53 | (1.08 - 2.18) | 1.43 | (1.00 - 2.03) | 1.55 | (1.09 - 2.21) |
| White + Black African | 33,629 | 11 | 4,531,702 | 0 | 0.3 | (0.16 - 0.54) | 1.34 | (0.74 - 2.41) | 1.22 | (0.67 - 2.20) | 1.33 | (0.74 - 2.41) | 1.33 | (0.73 - 2.40) |
| White + Asian | 34,679 | 14 | 4,531,702 | 0 | 0.38 | (0.22 - 0.64) | 1.56 | (0.92 - 2.63) | 1.5 | (0.89 - 2.53) | 1.51 | (0.89 - 2.55) | 1.44 | (0.85 - 2.44) |
| Other mixed | 64,567 | 21 | 4,531,702 | 0 | 0.29 | (0.19 - 0.45) | 1.24 | (0.80 - 1.90) | 1.16 | (0.75 - 1.78) | 1.21 | (0.79 - 1.85) | 1.23 | (0.80 - 1.89) |
| Indian | 436,692 | 357 | 11,460,760 | 0.03 | 0.7 | (0.63 - 0.78) | 1.38 | (1.24 - 1.54) | 1.27 | (1.13 - 1.41) | 1.24 | (1.11 - 1.39) | 1.28 | (1.14 - 1.44) |
| Pakistani | 302,940 | 214 | 7,952,710 | 0.03 | 0.6 | (0.52 - 0.69) | 1.67 | (1.45 - 1.91) | 1.36 | (1.18 - 1.56) | 1.15 | (1.00 - 1.32) | 1.06 | (0.92 - 1.22) |
| Bangladeshi | 70,396 | 49 | 1,848,344 | 0.03 | 0.6 | (0.45 - 0.79) | 2.21 | (1.67 - 2.93) | 1.8 | (1.36 - 2.39) | 1.43 | (1.07 - 1.89) | 1.35 | (1.02 - 1.80) |
| Other Asian | 224,309 | 151 | 5,889,946 | 0.03 | 0.59 | (0.50 - 0.69) | 1.76 | (1.49 - 2.07) | 1.6 | (1.36 - 1.89) | 1.54 | (1.30 - 1.82) | 1.56 | (1.32 - 1.85) |
| Caribbean | 83,419 | 164 | 2,184,796 | 0.08 | 1.61 | (1.37 - 1.88) | 1.54 | (1.31 - 1.80) | 1.31 | (1.12 - 1.54) | 1.19 | (1.02 - 1.40) | 1.45 | (1.24 - 1.70) |
| African | 192,521 | 100 | 5,056,550 | 0.02 | 0.45 | (0.37 - 0.54) | 2.05 | (1.68 - 2.50) | 1.75 | (1.43 - 2.14) | 1.73 | (1.41 - 2.11) | 1.78 | (1.46 - 2.18) |
| Other Black | 68,949 | 41 | 1,810,214 | 0.02 | 0.5 | (0.37 - 0.69) | 1.74 | (1.28 - 2.36) | 1.47 | (1.07 - 2.00) | 1.39 | (1.02 - 1.90) | 1.45 | (1.06 - 1.98) |
| Chinese | 105,447 | 28 | 2,770,299 | 0.01 | 0.25 | (0.17 - 0.36) | 0.99 | (0.68 - 1.43) | 0.96 | (0.66 - 1.39) | 1.11 | (0.76 - 1.60) | 1.22 | (0.84 - 1.77) |
| Other | 219,283 | 109 | 5,758,492 | 0.02 | 0.44 | (0.37 - 0.54) | 1.36 | (1.13 - 1.65) | 1.26 | (1.04 - 1.53) | 1.26 | (1.04 - 1.52) | 1.26 | (1.04 - 1.52) |
| Unknown | 4,602,822 | 3,967 | 120,574,720 | 0.03 | 0.86 | (0.83 - 0.89) | 0.87 | (0.84 - 0.91) | 0.88 | (0.85 - 0.91) | 0.95 | (0.92 - 0.99) | 1.01 | (0.97 - 1.05) |
| **Non-COVID related death** |  |  |  |  |  |  |  |  |  |  |  |  |  |  |
| British | 9,598,922 | 53,687 | 251,408,914 | 0.21 | 1 |  | 1 |  | 1 |  | 1 |  | 1 |  |
| Irish | 79,374 | 554 | 2,076,507 | 0.27 | 1.37 | (1.26 - 1.50) | 1.11 | (1.02 - 1.21) | 1.06 | (0.98 - 1.16) | 0.99 | (0.91 - 1.08) | 0.98 | (0.90 - 1.06) |
| Other White | 1,352,377 | 2,914 | 35,498,448 | 0.08 | 0.41 | (0.40 - 0.43) | 0.9 | (0.87 - 0.94) | 0.88 | (0.84 - 0.91) | 0.91 | (0.87 - 0.94) | 0.92 | (0.88 - 0.95) |
| White + Black Caribbean | 39,676 | 80 | 4,531,702 | 0.02 | 0.38 | (0.31 - 0.48) | 0.99 | (0.80 - 1.24) | 0.9 | (0.72 - 1.12) | 0.94 | (0.75 - 1.17) | 0.98 | (0.79 - 1.22) |
| White + Black African | 33,629 | 32 | 4,531,702 | 0.01 | 0.18 | (0.13 - 0.26) | 0.75 | (0.53 - 1.06) | 0.7 | (0.49 - 0.99) | 0.82 | (0.58 - 1.16) | 0.81 | (0.57 - 1.14) |
| White + Asian | 34,679 | 27 | 4,531,702 | 0.01 | 0.15 | (0.10 - 0.22) | 0.6 | (0.41 - 0.87) | 0.58 | (0.40 - 0.85) | 0.64 | (0.44 - 0.93) | 0.62 | (0.43 - 0.91) |

|  | Denomina<br>tor | Event | Total person-weeks | Rate per<br>1,000 | Crude | Age-sex adjusted | plus IMD | plus co-morbidities,<br>GP consults | plus household size<br>and carehome |
| --- | --- | --- | --- | --- | --- | --- | --- | --- | --- |
| Other mixed | 64,567 | 53 | 4,531,702 | 0.01 | 0.16 (0.12 - 0.21) | 0.64 (0.49 - 0.83) | 0.6 (0.45 - 0.78) | 0.66 (0.50 - 0.86) | 0.66 (0.50 - 0.86) |
| Indian | 436,692 | 891 | 11,460,760 | 0.08 | 0.4 (0.38 - 0.43) | 0.79 (0.74 - 0.85) | 0.74 (0.69 - 0.79) | 0.87 (0.81 - 0.93) | 0.89 (0.83 - 0.95) |
| Pakistani | 302,940 | 529 | 7,952,710 | 0.07 | 0.33 (0.30 - 0.36) | 0.88 (0.80 - 0.95) | 0.74 (0.67 - 0.80) | 0.77 (0.70 - 0.84) | 0.74 (0.68 - 0.81) |
| Bangladeshi | 70,396 | 93 | 1,848,344 | 0.05 | 0.25 (0.21 - 0.31) | 0.9 (0.73 - 1.10) | 0.76 (0.62 - 0.93) | 0.76 (0.62 - 0.93) | 0.74 (0.60 - 0.91) |
| Other Asian | 224,309 | 272 | 5,889,946 | 0.05 | 0.25 (0.22 - 0.28) | 0.72 (0.64 - 0.82) | 0.68 (0.60 - 0.76) | 0.77 (0.69 - 0.87) | 0.78 (0.70 - 0.89) |
| Caribbean | 83,419 | 389 | 2,184,796 | 0.18 | 0.92 (0.84 - 1.02) | 0.91 (0.82 - 1.01) | 0.79 (0.72 - 0.88) | 0.81 (0.74 - 0.90) | 0.88 (0.80 - 0.97) |
| African | 192,521 | 174 | 5,056,550 | 0.03 | 0.18 (0.15 - 0.21) | 0.76 (0.66 - 0.89) | 0.67 (0.57 - 0.77) | 0.79 (0.68 - 0.92) | 0.79 (0.68 - 0.92) |
| Other Black | 68,949 | 108 | 1,810,214 | 0.06 | 0.31 (0.25 - 0.37) | 0.98 (0.82 - 1.19) | 0.87 (0.71 - 1.05) | 0.9 (0.74 - 1.08) | 0.91 (0.75 - 1.10) |
| Chinese | 105,447 | 66 | 2,770,299 | 0.02 | 0.12 (0.10 - 0.15) | 0.46 (0.36 - 0.59) | 0.46 (0.36 - 0.58) | 0.59 (0.46 - 0.75) | 0.6 (0.47 - 0.77) |
| Other | 219,283 | 271 | 5,758,492 | 0.05 | 0.24 (0.22 - 0.28) | 0.74 (0.65 - 0.83) | 0.7 (0.62 - 0.79) | 0.74 (0.66 - 0.84) | 0.74 (0.66 - 0.84) |
| Unknown | 4,602,822 | 24,732 | 120,574,720 | 0.21 | 0.97 (0.95 - 0.98) | 0.96 (0.94 - 0.97) | 0.96 (0.95 - 0.98) | 1.06 (1.05 - 1.08) | 1.09 (1.08 - 1.11) |

**Figure S1.**
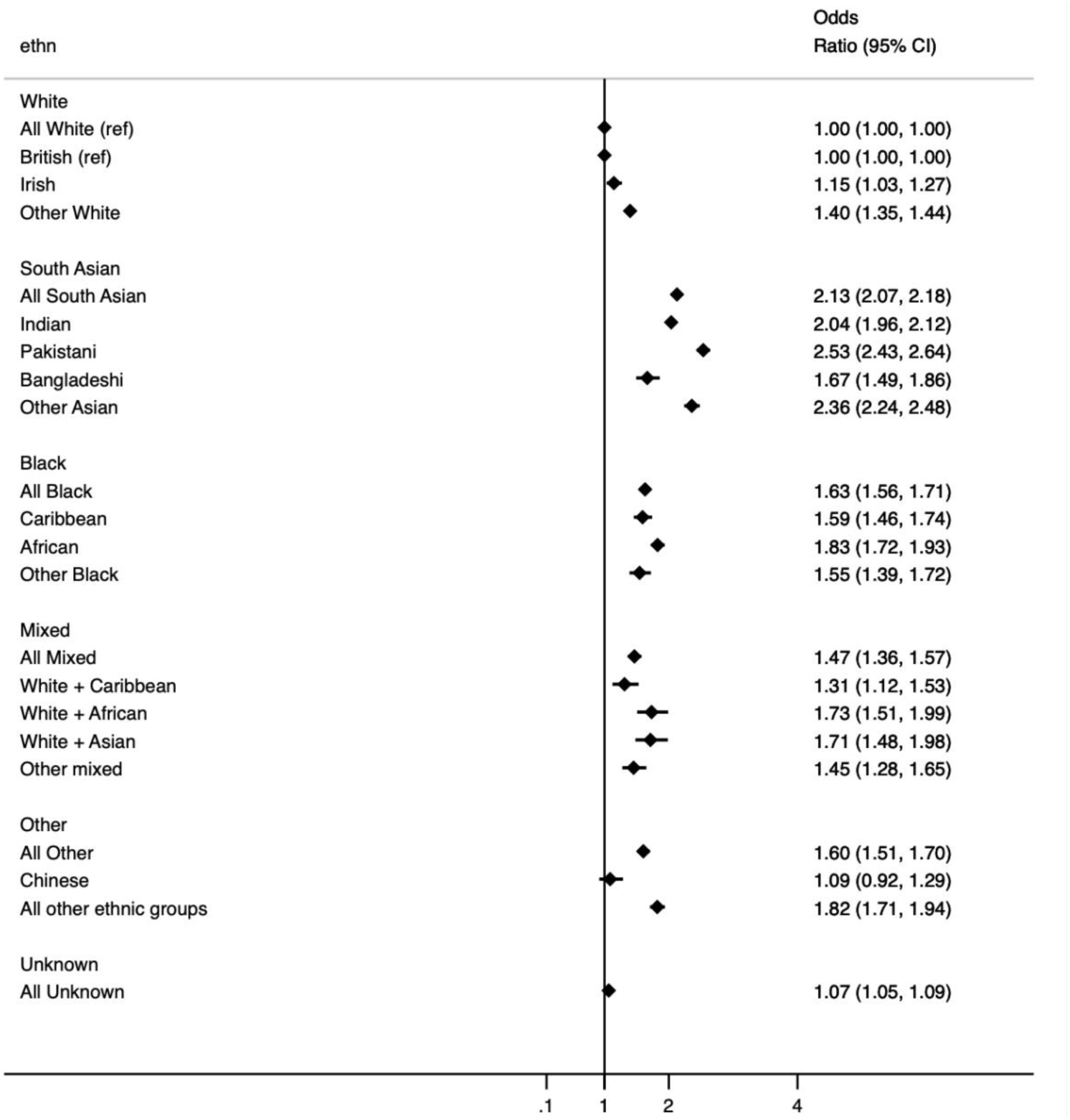
Odds of testing positive amongst those ever receiving a test. ∗All White is the reference category for comparison of ethnicity in 5 categories. British is the reference category for comparison of ethnicity in 16 categories. ∗models adjust for age, sex, deprivation quintile, all pre-specified clinical co-morbidities, categories of BMI, HbA1c, and systolic and diastolic blood pressure, number of primary care consultations in the 12 months prior, household size, care home residency, and stratification by STP region.

## References

1. Pan, D. et al.. The impact of ethnicity on clinical outcomes in COVID-19: A systematic review. EClinicalMedicine 23, 100404 (2020).

2. The OpenSAFELY Collaborative et al. OpenSAFELY: factors associated with COVID-19- related hospital death in the linked electronic health records of 17 million adult NHS patients. Epidemiology (2020) doi:10.1101/2020.05.06.20092999.

3. Aldridge, R. W. et al.. Black, Asian and Minority Ethnic groups in England are at increased risk of death from COVID-19 : indirect standardisation of NHS mortality data. 1–7 (2020).

4. Baumer, T. et al.. Insights into the Epidemiology of the First Wave of COVID-19 ICU Admissions in South Wales--the Interplay between Ethnicity and Deprivation. (2020).

5. Public Health England. Beyond the data: Understanding the impact of COVID-19 on BAME groups. https://assets.publishing.service.gov.uk/government/uploads/system/uploads/attachment_data/file/892376/COVID_stakeholder_engagement_synthesis_beyond_the_data.pdf (2020).

6. Baqui, P., Bica, I., Marra, V., Ercole, A. & van der Schaar, M. Ethnic and regional variations in hospital mortality from COVID-19 in Brazil: a cross-sectional observational study. Lancet Glob Health 8, e1018–e1026 (2020).

7. Rentsch CT, Kidwai-Khan F, Tate JP, Park LS, King JT, J., Skanderson M, et al. Patterns of COVID-19 testing and mortality by race and ethnicity among United States veterans: A nationwide cohort study. PLoS Med. 17, e1003379 (2020).

8. Lassale, C., Gaye, B., Hamer, M., Gale, C. R. & Batty, G. D. Ethnic disparities in hospitalisation for COVID-19 in England: The role of socioeconomic factors, mental health, and inflammatory and pro-inflammatory factors in a community-based cohort study. Brain Behav. Immun. (2020) doi:10.1016/j.bbi.2020.05.074.

9. Hawkins, D. Differential occupational risk for COVID‐19 and other infection exposure according to race and ethnicity. Am. J. Ind. Med. (2020) doi:10.1002/ajim.23145.

10. Rimmer, A. Covid-19: Disproportionate impact on ethnic minority healthcare workers will be explored by government. BMJ (2020) doi:10.1136/bmj.m1562.

11. Gupta, R., Hussain, A. & Misra, A. Diabetes and COVID-19: evidence, current status and unanswered research questions. Eur. J. Clin. Nutr. 74, 864–870 (2020).

12. Pareek, M. et al.. Ethnicity and COVID-19: an urgent public health research priority. Lancet 395, 1421–1422 (2020).

13. Dyson, M. COVID-19: the risk to BAME doctors. https://www.bma.org.uk/advice-and-support/covid-19/your-health/covid-19-the-risk-to-bame-doctors (2020).

14. Mahase, E. Covid-19: Ethnic minority doctors feel more pressured and less protected than white colleagues, survey finds. BMJ 369, (2020).

15. Race Relations (Amendment) Act 2000. (2000).

16. Hull, S. et al.. Research into practice: understanding ethnic differences in healthcare usage and outcomes in general practice. Br. J. Gen. Pract. 64, 653–655 (2014).

17. Mathur, R., Grundy, E. & Smeeth, L. Availability and use of UK based ethnicity data for health research. National Centre for Research Methods Working Paper Series 1–30 (2013).

18. Mathur, R. et al.. Completeness and usability of ethnicity data in UK-based primary care and hospital databases. J. Public Health 36, 684–692 (2014).

19. Sheldon, T. A., Parker, H., T.a., S. & H., P. Race and ethnicity in health research. J. Public Health Med. 14, 104–110 (1992).

20. Mathur, R. et al.. Is individual smoking behaviour influenced by area-level ethnic density? A cross-sectional electronic health database study of inner south-east London. ERS Monograph 3, (2017).

21. Garner, S. & Bhattacharyya, G. Poverty, ethnicity and place. York: Joseph Rowntree Foundation (2011).

22. Harrison, E. M. et al.. Ethnicity and Outcomes from COVID-19: The ISARIC CCP-UK Prospective Observational Cohort Study of Hospitalised Patients. (2020) doi:10.2139/ssrn.3618215.

23. Apea, V. J. et al.. Ethnicity and outcomes in patients hospitalised with COVID-19 infection in East London: an observational cohort study. medRxiv (2020).

24. Hull, S. A., Williams, C., Ashworth, M., Carvalho, C. & Boomla, K. Suspected COVID-19 in primary care: how GP records contribute to understanding differences in prevalence by ethnicity. medRxiv 2020.05.23.20101741 (2020).

25. Garg, S. et al.. Hospitalization Rates and Characteristics of Patients Hospitalized with Laboratory-Confirmed Coronavirus Disease 2019 — COVID-NET, 14 States, March 1–30, 2020. US Department of Health and Human Services/Center for Disease Control and Prevention (2020).

26. Arnold, D. T. et al.. Patient outcomes after hospitalisation with COVID-19 and implications for follow-up; results from a prospective UK cohort. Respiratory Medicine (2020) doi:10.1101/2020.08.12.20173526.

27. Vlachos, S. et al.. Hospital mortality and resource implications of hospitalisation with COVID-19 in London, UK: a prospective cohort study. Intensive Care and Critical Care Medicine (2020) doi:10.1101/2020.07.16.20155069.

28. Griffith, G. et al.. Collider bias undermines our understanding of COVID-19 disease risk and severity. Epidemiology (2020) doi:10.1101/2020.05.04.20090506.

29. Chris White And. Coronavirus (COVID-19) related deaths by ethnic group, England and Wales - Office for National Statistics. https://www.ons.gov.uk/peoplepopulationandcommunity/birthsdeathsandmarriages/deaths/articles/coronaviruscovid19relateddeathsbyethnicgroupenglandandwales/2march2020to15may2020 (2020).

30. ICNARC. ICNARC report on COVID-19 in critical care 31 July 2020. (2020).

31. OpenSAFELY Codelists: Ethnicity. https://codelists.opensafely.org/codelist/opensafely/ethnicity/2020-04-27/.

32. Rentsch, C. T. et al.. Hydroxychloroquine for prevention of COVID-19 mortality: a population-based cohort study. medRxiv 2020.09.04.20187781 (2020).

33. COVID-19 Racial Disparities in Testing, Infection, Hospitalization, and Death: Analysis of Epic Patient Data. https://www.kff.org/coronavirus-covid-19/issue-brief/covid-19-racial-disparities-testing-infection-hospitalization-death-analysis-epic-patient-data/ (2020).

34. Kenway, P. & Holden, J. Accounting for the Variation in the Confirmed Covid-19 Caseload across England: An analysis of the role of multi-generation households, London and time. New Policy Institute, London (2020).

35. Overcrowded households. https://www.ethnicity-facts-figures.service.gov.uk/housing/housing-conditions/overcrowded-households/latest (2020).

36. Platt, L. & Warwick, R. Are some ethnic groups more vulnerable to COVID-19 than others. Institute for Fiscal Studies, Nuffield Foundation (2020).

37. English language skills. https://www.ethnicity-facts-figures.service.gov.uk/ukpopulation-by-ethnicity/demographics/english-language-skills/latest (2018).

38. Stop the coronavirus stigma now. Nature 580, 165 (2020).

39. Dr Ben Windsor-Shellard And. Coronavirus (COVID-19) related deaths by occupation, England and Wales - Office for National Statistics. https://www.ons.gov.uk/peoplepopulationandcommunity/healthandsocialcare/causesofdeath/bulletins/coronaviruscovid19relateddeathsbyoccupationenglandandwales/deathsregisteredbetween9marchand25may2020 (2020).

40. Bhopal, R. S. et al.. Mortality, ethnicity, and country of birth on a national scale, 2001- 2013: A retrospective cohort (Scottish Health and Ethnicity Linkage Study). PLoS Med. 15, e1002515 (2018).

41. Scott, A. P. & Timæus, I. M. Mortality differentials 1991-2005 by self-reported ethnicity: findings from the ONS Longitudinal Study. J. Epidemiol. Community Health 67, 743–750 (2013).

42. Stevenson, O. & Morris, S. Coronavirus: record ethnicity on all death certificates to start building a clearer picture. The Conversation (2020).

43. Christie, B. Scotland introduces record of ethnicity on death certificates. BMJ 344, e475 (2012).

44. London Health Obsrevatory. Missing Record: The Case for Recording Ethnicity at Birth and Death Registrations. https://www.kent.ac.uk/chss/docs/Births_Deaths_Reg1.pdf (2003).

